# The Pathogen Adaptation of HLA Alleles and the Correlation with Autoimmune Diseases based on the HLA Diversity Resource in the Han Chinese

**DOI:** 10.1101/2024.11.19.24317546

**Authors:** Shuai Liu, Yanyan Li, Tingrui Song, Jingjing Zhang, Peng Zhang, Huaxia Luo, Sijia Zhang, Yiwei Niu, Tao Xu, Shunmin He

## Abstract

Human leukocyte antigen (HLA) genes play a crucial role in the adaptation of human populations to the dynamic pathogenic environment. Despite their significance, investigating the pathogen-driven evolution of HLAs and the implications for autoimmune diseases presents considerable challenges. Here, we genotyped over twenty HLA genes at 3-field resolution in 8278 individuals from diverse ethnic backgrounds, including 4013 unrelated Han Chinese. We focused on the adaptation of HLAs in the Han Chinese by analysing their binding affinity for various pathogens, and explored the potential correlations between pathogen adaptation and autoimmune diseases. Our findings reveal that specific HLA alleles like HLA-DRB1*07:01 and HLA-DQB1*06:01, confer strong pathogen adaptability at the sequence level, notably for *Corynebacterium diphtheriae* and *Bordetella pertussis*. Additionally, alleles like HLA*03:02 demonstrate adaptive selection against pathogens like *Mycobacterium tuberculosis* and *Coronavirus* at the gene expression level. Simultaneously, the aforementioned HLA alleles are closely related to some autoimmune diseases such as multiple sclerosis (MS). These exploratory discoveries shed light on the intricate coevolutionary relationships between pathogen adaptation and autoimmune diseases in the human population. These efforts led to an HLA database at http://bigdata.ibp.ac.cn/HLAtyping, aiding searches for HLA allele frequencies (AF) across populations.

## Introduction

The immune process is of paramount importance in the intricate interplay between the *in vivo* life system and the surrounding pathogenic microbial environment [1]. The HLA genes located on chromosomal region 6p21.3, in particular the classical HLA genes (*HLA-A*, *HLA-B*, *HLA-C*, *HLA-DRB1*, *HLA-DPA1*, *HLA-DPB1*, *HLA-DQA1*, *HLA-DQB1*), play a pivotal role in the identification and presentation of invasive foreign pathogens to the immune system [2]. As a result, these classical HLA genes are under intense selective pressure for resistance against pathogens, resulting in the highest degree of polymorphism observed in the human genome [3,4]. To date, two primary mechanistic theories, negative frequency-dependent selection (NFDS) and heterozygote advantage (HA), have been proposed to explain the remarkable diversity of HLA genes in the fluctuating pathogenic microbial environment [5]. Genetic predisposition to different pathogens is an area of increasing interest [6]. Although many human HLA resources have been developed [7–14], there are few studies disclosing the HLA frequency database of the Han Chinese population with high resolution at the exon level and focusing on human pathogen co-evolution. By exploiting the binding affinity of HLA molecules for pathogenic peptides, we can gain valuable insights into the evolutionary history of human adaptation to pathogens [15,16].

There has always been a phenomenon of co-adaptation between humans and pathogens, which has had an important impact on many human diseases, a typical example being autoimmune diseases. The hypothesis of “pathogen-driven selection” provides important insights into the mechanisms of the evolution of autoimmune diseases [17,18]. An ancient genomics study shows that immunity genes have been strongly affected by both positive and negative selection, and that resistance to infection has increased the risk of inflammatory disease over the past millennia [19]. Despite this, HLA genes are the most important genes for pathogen adaptation and autoimmunity, and there are few studies based on them to elucidate the genetic impact of pathogen adaptation on autoimmune disease in the population.

In this study, we performed high-resolution genotyping of HLA alleles in a large cohort consisting of 4129 unrelated individuals from the NyuWa Genome Project [20], 3202 unrelated individuals from the 1000 Genomes Project (1KGP) [21], and 893 unrelated individuals from the Human Genome Diversity Project (HGDP) [22]. Our comprehensive analysis extended to characterising the binding affinity of common HLA alleles in the Han Chinese population for a range of human epidemic pathogens, and evaluating the influence of pathogen adaptation of HLA alleles on autoimmune disease. Finally, we explored the adaptive selection on the transcriptional regulation of the HLA alleles under the pathogen pressure. The analysis of HLA alleles in a large Han population helps us to understand the adaptation landscape of populations to pathogens and the relationship between pathogen adaptation and autoimmune disease.

## Results

### High-resolution Genotyping of HLA Genes in Multiracial Populations

In this comprehensive study, we successfully genotyped a total of 31 HLA genes. Our analysis was grounded in the assessment of genotyping rate, resolution, and accuracy across 8278 samples from the NyuWa project, the 1KGP, and the HGDP. The genotyping rates of various HLA genes within these cohorts revealed that the classical HLA genes, including *HLA-A*, *HLA-B*, *HLA-C*, *HLA-DRB1*, *HLA-DQA1*, *HLA-DQB1*, *HLA-DPA1*, and *HLA-DPB1*, were genotyped in all samples (Figure 1A). Moreover, all these HLA genes achieved a minimum of 2-field genotyping level (amino acid resolution), with majority achieving the 3-field genotyping level (exon sequence resolution) (Figure 1A). Upon comparing our genotyping results with benchmark genotypes for *HLA-A*, *HLA-B*, *HLA-C*, *HLA-DRB1*, and *HLA-DQB1* from the 1KGP, we observed that the genotyping accuracy for these genes exceeded 99% at the 1-field genotyping level (serotyping level), and had an impressive 94% to 97% accuracy at the amino acid level. (Figure 1B). In addition, we quantified the genetic diversity by calculating the heterozygosity of HLA genes with genotyping rates surpassing 80%. The findings indicate that the seven classic HLA genes, *HLA-A*, *HLA-B*, *HLA-C*, *HLA-DRB1*, *HLA-DQA1*, *HLA-DQB1*, and *HLA-DPB1*, exhibit exceptionally high heterozygosity, with values exceeding 80% within the population (Figure 1C). This high level of heterozygosity underscores their critical role in the adaptation to pathogens [3,4]. The genetic architecture of HLA genes in the Han Chinese closely resembles that of East Asian population but exhibits significant differences when compared to European and African populations, in particular, the most frequent HLA haplotype (HLA-A*02:07–HLA-C*01:02–HLA-B*46:01–HLA-DRA*01:01–HLA-DRB1*09:0 1–HLA-DQA1*03:02–HLA-DQB1*03:03–HLA-DPA1*02:02–HLA-DPB1*05:01) in the Han Chinese only accounted for 0.19%, while the most frequent HLA haplotype (HLA-A*02:07–HLA-C*01:02–HLA-B*46:01–HLA-DRA*01:01–HLA-DRB1*14:5 4–HLA-DQA1*01:04–HLA-DQB1*05:02–HLA-DPA1*02:02–HLA-DPB1*02:02) in the East Asian accounted for 0.30%, and the most frequent HLA haplotype (HLA-A*03:01–HLA-C*07:02–HLA-B*07:02–HLA-DRA*01:02–HLA-DRB1*15:0 1–HLA-DQA1*01:02–HLA-DQB1*06:02–HLA-DPA1*01:03–HLA-DPB1*04:01) in the European accounted for 0.40%, and the most frequent HLA haplotype (HLA-A*30:01–HLA-C*17:01–HLA-B*42:01–HLA-DRA*01:02–HLA-DRB1*03:0 2–HLA-DQA1*04:01–HLA-DQB1*04:02–HLA-DPA1*02:02–HLA-DPB1*01:01) in the African accounted for 0.23% (Figure S1A–D).

**Figure 1.**
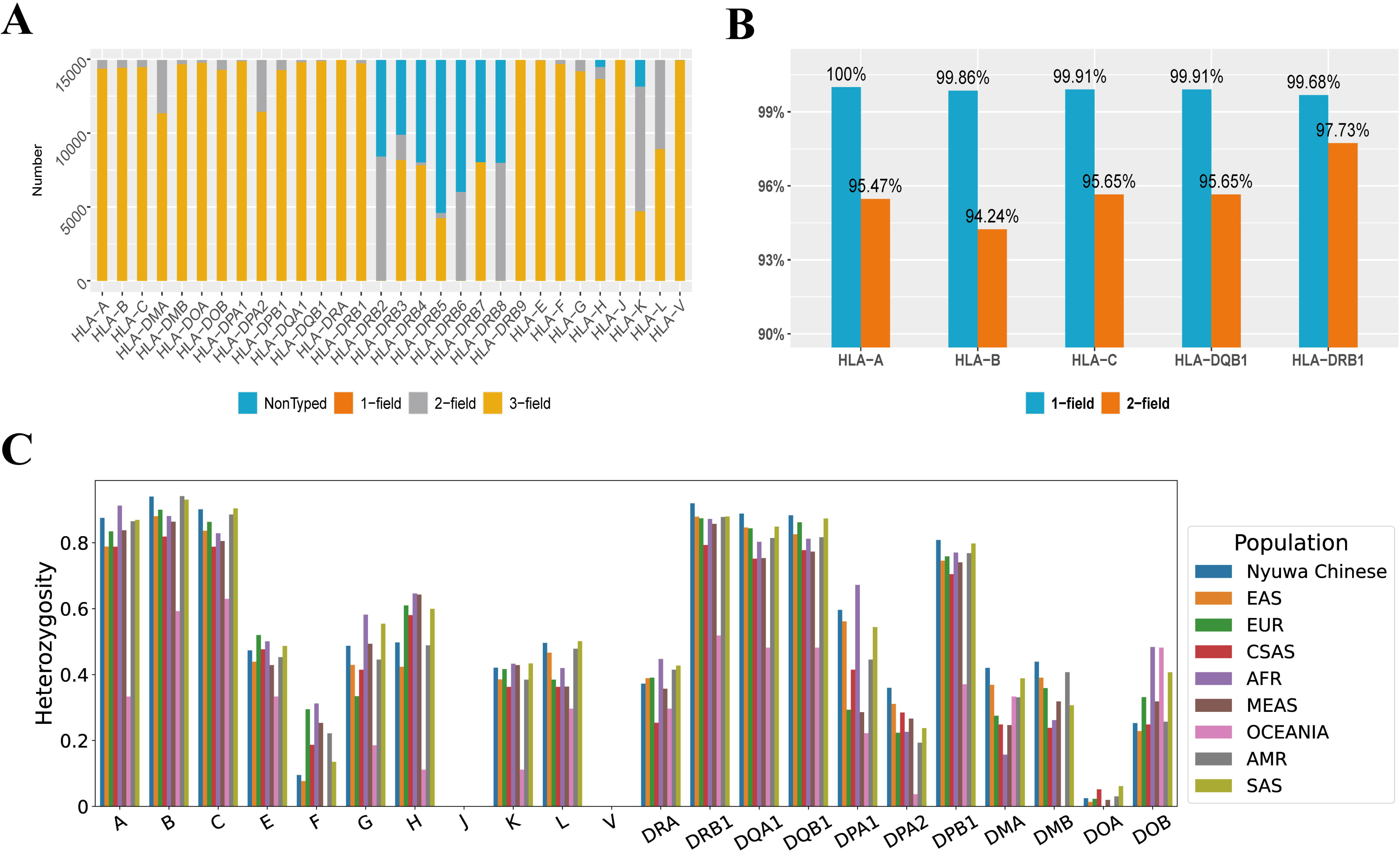
The evaluation and description for genotyped HLA genes. **A.** Proportion of individuals with completed HLA genotyping and the resolution of genotyped HLA alleles. The designation “not typed” signifies that the NGS reads of an individual could not be identified as any known HLA allele by HLA-HD software. **B.** The accuracy of HLA genotyping based on HLA-HD. This evaluation was performed based on the SBT-sequenced benchmark HLA genotypes from 1KGP. **C.** the heterozygosity of HLA genes in different populations.

Compared with the Allele Frequency Net Database [23], the HLA genes resource in this study provides HLA alleles of classification precision up to 3-field of large-scale of Han Chinese individuals, as well as multi-ethnic populations around the world. HLA alleles with 2-field or higher genotyping fields can provide information on the amino acid chain, which could help us complete molecular docking to determine the ability of HLA molecules to recognise foreign substances, and further find out the recent adaptation landscapes of HLA alleles in human populations. In addition, this HLA data resource has filled many missing genotypes of HLA genes in the northern Han population, providing a more complete perspective to understand the diversity and adaptive evolution of HLA genes in the Han Chinese population.

### Pathogen-peptide-binding Landscape of HLA Elements in the Han Chinese Population

In order to ascertain potential correlations between HLA-peptide binding affinity and pathogen adaptation, an investigation was conducted into the impact of HLA peptide binding affinity on the adaptability of the human immune system in the context of HIV and HCV infection. This investigation focused in particular on the associated HLA alleles as reported in studies [24,25]. The results indicate a significant correlation between an increased HLA binding affinity to HIV or HCV peptides and a reduced capacity for pathogen invasion and persistence (Figure S2, S3). These findings reveal a critical aspect of HLA adaptation to these viruses, namely that a higher affinity confers an enhanced ability to identify and combat these pathogens. Although HIV and HCV have only been present in the human population for only a few decades, the population’s adaptation to the consensus sequences of other pathogens can lead to the adaptation of HIV and HCV [26,27]. This discovery has profound biological significance, as it underscores the strategic use of affinity scores of HLA molecules for pathogen-derived peptides in the prediction and analysis of immune responses, thereby enhancing our understanding of host-pathogen interaction and co-evolution.

By evaluating the binding affinity of both the common HLA types (AF >= 5%) and the low-frequency HLA types (AF >= 0.01 and AF < 0.05) with various epidemic pathogens (Table S1), we observed that the different alleles of *HLA-DRB1*, regardless of their AF, demonstrated a stronger ability to bind to a wide range of pathogen antigens (Figure 2A and Figure 2B). This phenomenon is also observed for the rare *HLA-DRB1* alleles in the population (Figure S4), as well as for *HLA-DRB1* alleles in other populations (Figure S5 and Figure S6). Specifically, HLA-DRB1*07:01 (AF = 8.1%) shows a robust binding affinity to *Corynebacterium diphtheriae*, and HLA-DRB1*08:03 (AF = 6.2%) exhibits a strong binding affinity to *Clostridium tetani* and *Bacillus anthracis* (Figure 2A). Additionally, HLA-DRB1*14:54 (AF = 2.7%) displays a notable binding affinity to *Bacillus anthracis* (Figure 2B). It is important to note that there is still a subtle tendency for the adaptive HLA types to bind to peptides. The most common peptide type bound by HLA-DRB1*07:01 is “LIVS/T/KALKLI/L” (Figure S7A), while the most common peptide type bound by HLA-DRB1*08:03 is “YV/KSSI/KK/NKILD” (Figure S7B), and “YL/II/KK/IKKNIE” is the most common peptide type bound by HLA-DRB1*12:02 (Figure S7C), and “IKI/NS/EK/SKE/NLL/I” is the most common peptide type bound by HLA-DRB1*14:54 (Figure S7D). This preference for HLA peptide affinity suggests that adaptive HLA types may only target one of the prevalent pathogens with the preferred peptide, and adaptation to the rest of the pathogens is simply a result of passive adaptation to a pathogen that also carries the preferred peptide. Furthermore, HLA-DQB1*03:01 and HLA-DQB1*06:01 show a pronounced affinity for specific extracellular pathogens, such as *Mycobacterium tuberculosis* and *Bordetella pertussis* (Figure 2A). The affinity of the two *HLA-DQB1* alleles for the three extracellular pathogen antigenic peptides also showed a clear preference for a particular type of amino acid sequence (Figure S8). Among the peptides bound by HLA-DQB1*03:01, there was a clear preference for the binding of the “VAAAAAAAA” peptide type (Figure S8A), whereas among the peptides bound by HLA-DQB1*06:01, the “VAAAAAAAAA” peptide type was also significantly preferred for binding (Figure S8B). This result suggests that the two HLA alleles that appear to be adapted to one of these three pathogens are also adapted to the other two.

**Figure 2.**
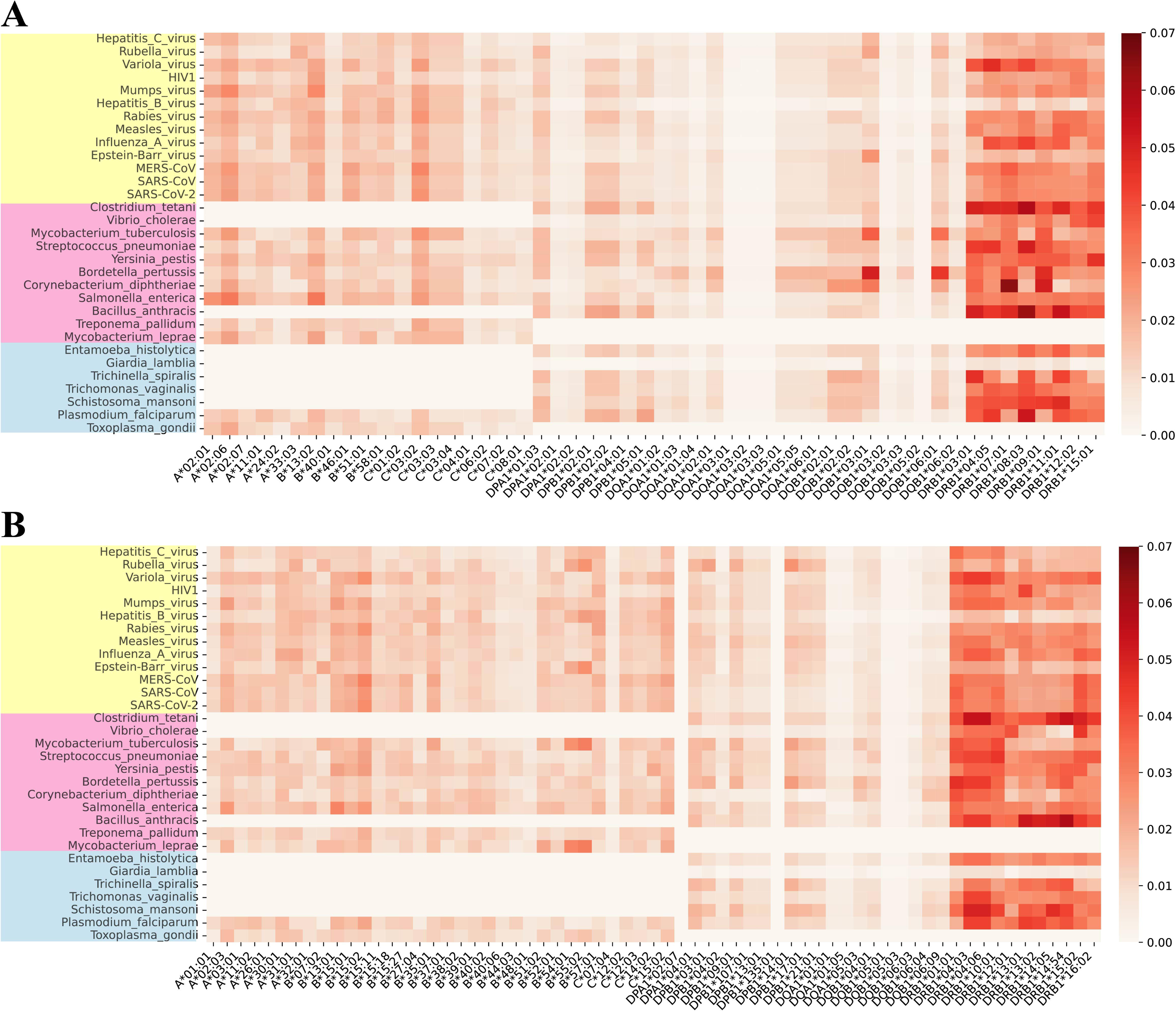
Binding affinity between common and low-frequency HLAs in the Han Chinese and the epidemic pathogens. **A.** Common HLAs**. B.** Low-frequency HLAs. The pathogens represented by the yellow block are viruses, those represented by the pink block are bacteria, and those represented by the blue block are parasites. A higher score indicates a greater affinity.

### Antagonistic Interactions between HLA Alleles involved in Adaptation and Autoimmune

In order to investigate the genetic correlations between autoimmune diseases and pathogen adaptation in the human population, a correlation analysis was performed on the HLA alleles in the Han Chinese population and the autoimmune diseases related HLA alleles (Table S2). In general, a significant proportion of these *HLA-DRB1* alleles (HLA-DRB1*03:01, HLA-DRB1*04:05, HLA-DRB1*07:01, HLA-DRB1*09:01, HLA-DRB1*01:01, HLA-DRB1*13:01, HLA-DRB1*13:02) and the HLA-DQB1*06:01, which confers resistance to pathogens, has also been identified as the susceptibility alleles for numerous autoimmune diseases (Figure 3, Figure 4 and Figure S9). The pleiotropy of these HLA alleles provides an antagonistic balance between protection against foreign pathogens and the risk of autoimmunity [19]. In comparison to other ethnic groups, the HLA-DRB1*07:01 and HLA-DRB1*13:01 alleles in European and African populations exert a pleiotropic effect on the correlation between susceptibility to autoimmune diseases and pathogen adaptation (Figure S10 and Figure S11). Moreover, the high genetic linkage of HLA genes means that many pathogen-adapted HLA alleles also influence the population frequencies of autoimmune-related HLA alleles [28]. Indeed, it has been observed that over 80% of these common HLA alleles (Figure 3) and half of these low-frequency HLA alleles (Figure 4) show a significant genetic linkage to autoimmune-related HLA alleles in the Han Chinese population. In addition to the pleiotropic effects of HLA alleles, the high genetic linkage between HLA alleles represents a further significant factor contributing to the antagonistic balance between pathogen adaptation and disease risk. To illustrate, HLA-DRB1*07:01 exhibits a robust binding affinity for the antigen of *Corynebacterium diphtheriae*, and it also shows significantly positive correlations with HLA-DQB1*02:02 (Pearson correlation coefficient *r*^2^ = 0.72; the genetic distance between two HLA alleles *DQB1_CM_* – *DRB1_CM_* = 0.058cM) and HLA-DQA1*02:01 (*r*^2^ = 0.99; *DRB1_CM_* – *DQA1_CM_* = 0.014cM). The former is a risk factor for coeliac disease (CD) [29], while the latter is associated with an increased risk of inflammatory bowel disease (IBD) [30] (Figure 3).

**Figure 3.**
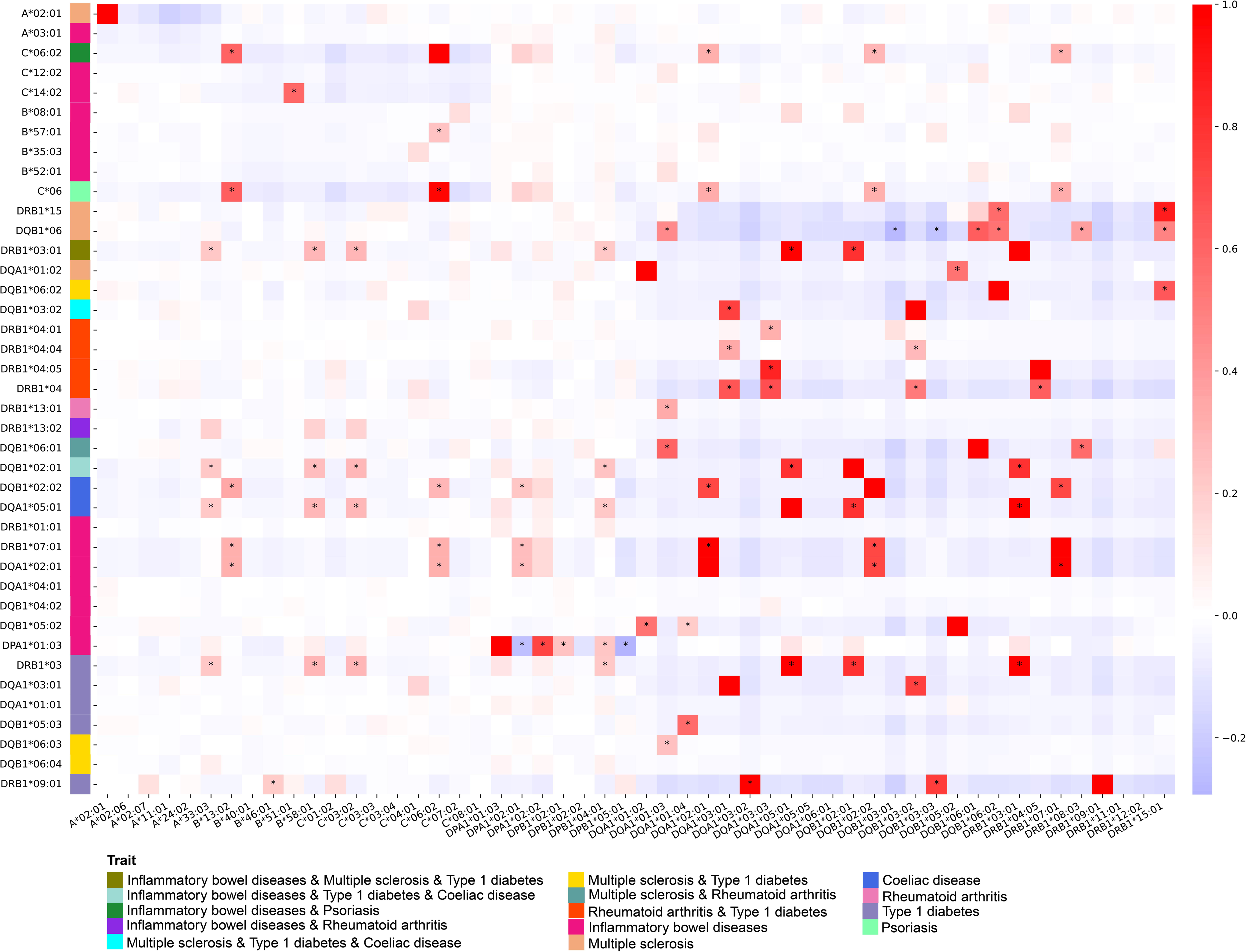
Associations between the common HLA alleles in the Han Chinese and the autoimmune susceptibility HLA alleles. The horizontal axis represents common potential adaptive HLA alleles in the Han Chinese population, and the vertical axis represents autoimmune susceptibility HLA alleles. Pearson correlation tests were performed on the genotypes of the two sets of allele sets, and “*” indicates a significantly correlated gene pair (r^2^ > 0.2 and P < 0.05).

**Figure 4.**
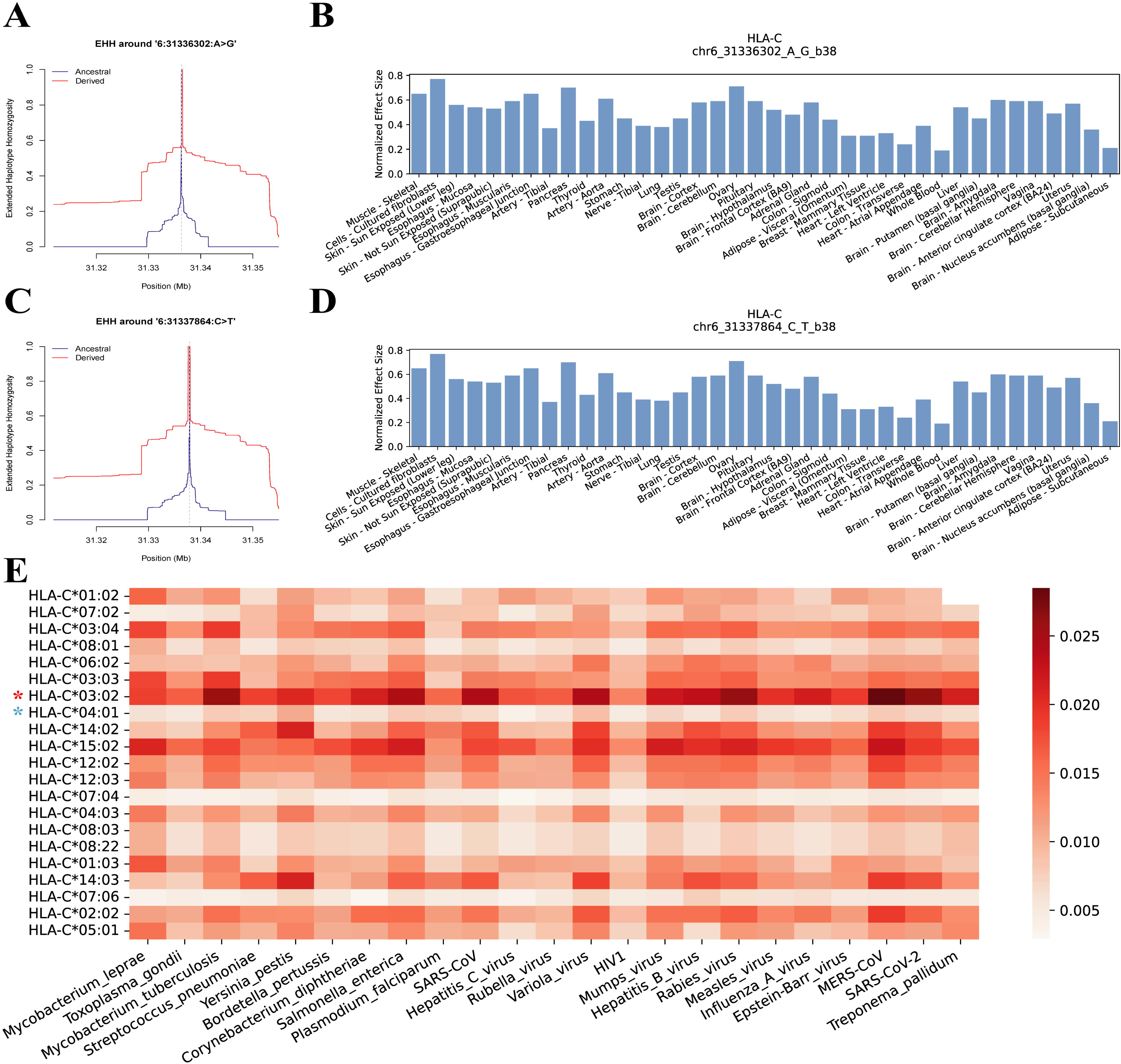
The transcriptional regulation of *HLA-C* under recent positive selection. **A.** The EHH plot of the focal marker 6:31336302A>G. **B.** The normalized effect size of 6:31336302A>G on the expression of HLA-C in tissues. **C.** EHH plot of the focal marker 6:31337864C>T. **D.** The normalized effect size of 6:31337864C>T on the expression of *HLA-C* in tissues. **E.** The binding affinity of *HLA-C* alleles with 20 pathogens that secretes intracellular antigens. A higher score indicates a greater binding affinity. It is notable that the *HLA-C* allele marked with a red star represents a significant positive correlation with the favoured regulator, while the *HLA-C* allele marked with a blue star represents a significant negative correlation with the favoured regulator.

Additionally, there are complex scenarios in which HLA alleles that interact with one another exhibit varying degrees of adaptability to different pathogens. Collectively, they influence the prevalence of autoimmune-related HLA alleles in diverse directions within the population. To illustrate, HLA-DQB1*06 is a risk factor for MS [31], and has a strong positive correlation with many HLA alleles associated with pathogen adaptation, including some subtypes such as HLA-DQB1*06:01 etc., as well as the genetically linked HLA-DRB1*08:03 (*r*^2^ = 0.38), and HLA-DRB1*15:01 (*r*^2^ = 0.49; of the strongest susceptibility effect in MS [32]), also contains a pathogen-adapted HLA allele HLA-DQB1*03:01 (*r*^2^ = –0.28) with a significant negative correlation (Figure 3). *Clostridium tetani* which produces tetanus toxin, and *Bordetella pertussis* which produces pertussis toxin, are significant inducers of MS [33,34]. Therefore, we postulated that the *Clostridium tetani*, which had left adaptive genetic signatures on HLA-DRB1*08:03, and the *Bordetella pertussis*, which had left adaptive genetic signatures on HLA-DQB1*03:01 (Figure 2A), may have a significant impact on the genetic susceptibility to MS in the Han Chinese population in the recent evolutionary history. As an additional example, HLA-DPB1*02:01, HLA-DPB1*04:01 and HLA-DPB1*05:01 represent the alleles at the *HLA-DPB1* locus, where the first two are positively correlated with HLA-DPA1*01:03, while the latter is negatively correlated with HLA-DPA1*01:03, which is the protective factor for IBD [30] (Figure 3). IBD has been reported to be associated with infection with a range of intestinal pathogens, including *Salmonella enterica*, *Epstein–Barr virus*, *Measles virus*, *Mumps virus*, *Rubella virus*, *Entamoeba histolytica*, *Toxoplasma gondii* etc. [35]. From a certain perspective, IBD has a more complex genetic structure due to the antagonistic evolution of HLA alleles and a wide range of intestinal pathogens.

### Pathogen-driven Adaptation of Transcriptional Regulation of *HLA-C*

In our work, we identified the upstream regulatory region of the *HLA-C* gene, which contains two genetically linked expression quantitative trait loci (eQTLs), 6: 31336302A>G and 6:31337864C>T, as a target of recent positive selection based on integrated haplotype score (iHS) and singleton density score (SDS) analyses (Figure S12; Figure S13). The derived alleles of these selected eQTLs exhibit a significantly extended haplotype homozygosity (Figure 4A and Figure 4C) and are characterised by a higher frequency in the East Asian population compared to the European and African populations (Figure S14). By making use of the resources on tissue-specific gene expression provided by the GTEx project, we were able to ascertain that the favoured alleles in question are prone to promoting the expression of *HLA-C* across a range of tissues (Figure 4B and Figure 4D). This elevated immune-related transcriptional level of HLA-C plays an essential role in strengthening the body’s defences against pathogens. In accordance with Ohta’s near-neutral theory of evolution, a mutation is considered nearly neutral if the selection coefficient *s* for the mutation meets the condition 0.2 < |2Ns| < 4 [36]. By tracking historical allele frequencies, it was observed that these two adaptive loci have been subject to a selection coefficient of 0.0009 (2 x Ne x s = 2 x 20000 x 0.0009 = 36 > 1, indicating a slightly strong selection pressure) (Figure S15; Figure S16). It is noteworthy that the eQTL 6:31337864C>T has been identified as a risk genetic factor for schizophrenia, with the potential for elevated prevalence due to the pressures of pathogen-driven selection.

Furthermore, the impact of recent positive selection on the regulation of *HLA-C* alleles was examined, and it was determined that HLA-C*03:02 and HLA-C*04:01 are significantly linked with favoured the eQTL (Figure 4E). HLA-C*03:02, which has a higher allele frequency in East Asians, has been associated with a broad and potent resistance to various pathogens, especially *Mycobacterium tuberculosis*, *Salmonella enterica*, *Variola virus*, *Rabies virus*, and *Coronavirus*. In contrast, HLA-C*04:01, which has the lowest allele frequency in East Asians, exhibits minimal resistance to pathogens (Figure 4E). Our findings suggest that the transcriptional regulation adjustment of HLA-C*03:02 has contributed to the enhanced general adaptability of the Han Chinese population to a range of pathogens. The affinity of HLA-C*03:02 for different pathogens is significantly different, which may be related to the specific preference of this allele for antigenic sequences. Therefore, we analysed the motif characteristics of peptides binding to HLA-C*03:02 and found that the amino acid fragments that can be bound by this HLA allele are very diverse and do not show significant motif preferences (Figure S17), which reveals the broad spectrum of adaptation of this HLA allele to pathogens.

It is also possible that *HLA-C*’s adaptation to pathogens at the gene expression level may affect HLA alleles related to human autoimmune diseases. Although HLA-C*04:01 does not show significant correlation with known autoimmune diseases related HLA alleles, HLA-C*03:02 makes a difference on four autoimmune diseases, including IBD, MS, Type 1 diabetes (T1D) and CD, in a way of genetic linkage (Figure 3). Here involved in four HLA alleles which have a significant positive correlation with HLA-C*03:02, consist of HLA-DRB*03:01 (*r*^2^ = 0.29; susceptible to MS and T1D, protective for IBD), HLA-DQB*02:01 (*r*^2^ = 0.28; susceptible to CD and T1D, protective for IBD), HLA-DQA1*05:01 (*r*^2^ = 0.29; susceptible to CD) and HLA-DRB*03 (*r*^2^ = 0.29; susceptible to T1D) (Figure 3 and Table S2). Therefore, from the perspective of the highly linked nature of the HLA region, the adaptive selection of HLA alleles in gene expression will also affect the prevalence of autoimmune diseases in the population.

### A High-resolution HLA Allele Frequency Database

In an effort to facilitate data sharing, we have established a comprehensive HLA allele frequency database (http://bigdata.ibp.ac.cn/HLAtyping) that enables users to search for allele frequencies and homozygosity of HLA genes across various populations, ranging from serotyping-level to exon-genotyping-level detail. This database offers a user-friendly interface with an intuitive search function on its homepage (Figure 5A), whereby users can input the name of the HLA allele to access relevant information. To further enhance the search experience, the database has been designed to include customizable options for selecting the genotyping fields (Figure 5B) and specifying the population of interest (Figure 5C). The database provides a comprehensive range of information for each HLA allele, including allele frequency, homozygosity and heterozygosity (proportion of carriers but not homozygotes) (Figure 5D). The allele frequency is a fundamental characteristic of HLA alleles within a population and is instrumental for analysing population genetic structure, inferring demographic history, and assessing the immune adaptability to pathogenic antigens (both infectious and tumourous antigens) among human populations. The proportion of homozygous and heterozygous forms of an HLA allele provides a more detailed understanding than allele frequency alone, offering insights into the individual-level adaptation to the pathogenic environments.

**Figure 5.**
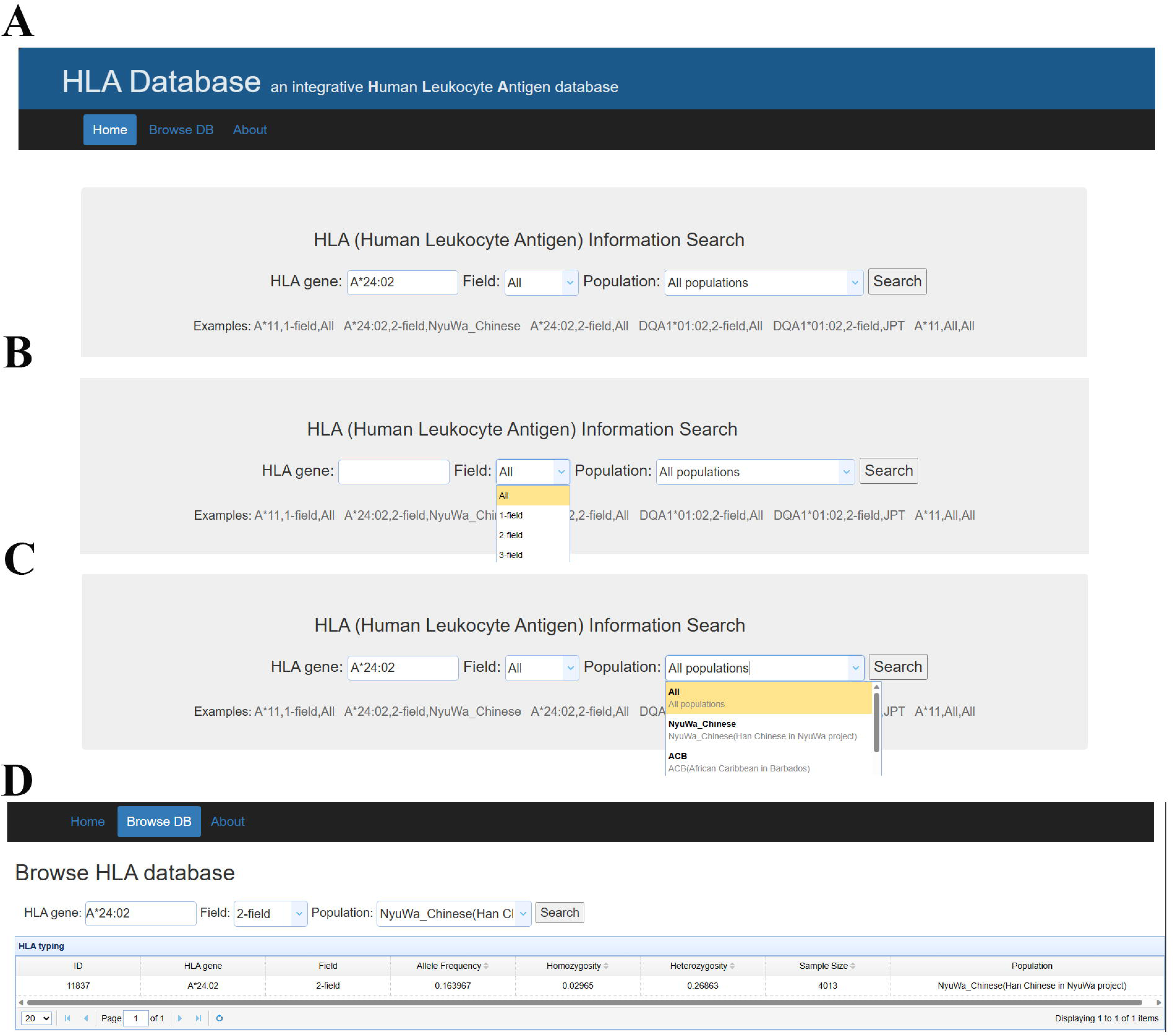
The graphic user interface of the HLA Database. **A.** A search bar is provided for the purpose of facilitating the input of search terms. **B.** The option for the field of HLA genotypes. **C.** The option for populations. **D.** Display of search results.

## Discussion

Pathogenic microbes represent an important driving force influencing human adaptive evolution [6]. It has been widely reported that HLA alleles are associated with susceptibility to pathogens [37–42] and autoimmune diseases [18,43–47]. The theory of pathogen-driven HLA diversity has been well developed to explain the significant association between the variants of HLA alleles and infectious diseases, as well as the autoimmune diseases [18,40,48]. Fortunately, the recent availability of large-scale of genome sequences in the human population [20–22] has facilitated the investigation of evolutionary relationships between HLA and pathogens at the population level. In light of the findings on host-pathogen co-evolution on MHC-peptide binding affinity from the studies [15,16], we performed an analysis of the binding affinity between pathogen peptides and the predominant HLA types in the Han Chinese population. Our results indicate that a significant number of HLA types possess varying degrees of binding ability to numerous pathogens (Figure 2), and are closely associated with the genetic risk of autoimmune diseases due to gene pleiotropy or genetic linkage (Figure 3).

The alleles of *HLA-DRB1* are found a notably stronger binding affinity to various pathogens than that of other HLA genes (Figure 2). This suggests that *HLA-DRB1* alleles have played a more substantial role in shaping the historical adaptation of human population to the intricate and varied pathogenic environment. *HLA-DRB1* plays a central role in the immune system by presenting peptides derived from extracellular proteins [49], and has the most diversity in the class II HLA genes [50] A phylogenetic study found that the *HLA-DRB1* gene clusters human-specific alleles [51], thus we could speculated that *HLA-DRB1* has been in a long-term arms race with various pathogens and may have contributed to lots of local adaptation of humans. Furthermore, in this paper, the affinity between HLA and pathogens is measured by the average of the affinity scores between HLA and different peptides of pathogen, which describes the relative probability score of an HLA molecule recognizing a pathogen antigen. Although the strong binding effect of specific antigenic peptides may be masked, for example, one peptide affinity is 0.85 from a pathogen, and three peptides with 0.30 for another pathogen and all of them is 0.9, there is currently no more suitable method.

Assessing the impact of pathogen adaptation on the prevalence of autoimmune diseases in human populations is an important issue. There are two major genetic factors mediating the interaction between pathogen adaptation and autoimmune diseases. One is the pleiotropy of HLA alleles. In the Finnish population, several HLA alleles have been reported to be associated with susceptibility to infectious diseases and autoimmune diseases, such as HLA-DQA1*03:01 and HLA-DQB1*03:02 [52]. Another one is the genetic linkage between nearby HLA alleles. Many of HLA-loci based associations result from linkage disequilibrium between the HLA gene studied and other HLA genes or non-HLA genes close by [53]. In this work, the gene pleiotropy and the genetic linkage between HLA genes represent a further avenue of enquiry in our research on measuring associations between the pathogen-adapted HLA alleles and the autoimmune-related HLA alleles. However, autoimmune diseases are also the interactive result of genes and living environment, and involve confounder factors such as sex bias and infections [54]. Therefore, it is still a challenge for us to discriminate the effects of HLA alleles on the induction of autoimmunity, especially when the HLA alleles are affected by pathogen adaptation, it is more difficult for us to quantify how the effect of specific pathogens on HLA genes will affect the occurrence and development of autoimmune diseases.

While our research, bolstered by clinical data on HIV and HCV infections [24,25], has identified a significant positive correlation between HLA-peptides binding affinity and the host’s susceptibility to infection, further clinical data encompassing a broader spectrum of pathogens is necessary to fully elucidate the extent of this association. Additionally, it is important to recognize the variability in pathogen strains across different geographical regions [55–57]. Consequently, future research endeavours should prioritize the investigation of human adaptive evolution to pathogens at the strain level, taking into account the diverse nature of these pathogens and their impact on human populations.

The topic of T cell clonality is also worthy of discussion. Following the processing of extracellular antigens into peptides and subsequent complexation with surface class II MHC molecules in on professional antigen-presenting cells such as dendritic cells, these MHC-peptide complexes are presented and recognised by CD4 helper-T-cells [58]. The clonotype of a T cell population represents a molecular description of the unique sequences required to produce the T cell’s TCR antigen specificity, as well as the specific V and J genes involved in the composite rearrangements, following the completion of the selection and maturation process of T cells in the thymus [59]. This adaptive immune process represents a pivotal step in the host’s recognition of exogenous pathogens. Future research may take this into account, as well as other immune processes involved in the recognition of exogenous pathogens.

## Materials and methods

### Data Resource Description

In our study, we utilised genome data from a total of 8278 human individuals for HLA genotyping. The dataset comprised 4013 unrelated individuals from the NyuWa Chinese project, 2504 unrelated individuals from the 1000 Genomes Project (1KGP) (https://www.internationalgenome.org/), and 828 unrelated individuals from the Human Genome Diversity Project (HGDP) (https://www.internationalgenome.org/), as well as 116, 698, 119 related samples in each genome project. The quality-controlled alignment files (BAM format) for the 1KGP and HGDP samples were downloaded from their respective repositories, and the whole genome sequencing data for the NyuWa project samples were processed in alignment with the methods described in the published paper[20]. Subsequently, all reads that had been assigned to the HLA region (chr6:28,510,120-33,480,577, according to the GRCh38 assembly) were extracted for HLA genotyping. This encompasses reads that correspond to HLA genes, as well as those in an unmapped state. This approach ensures the full utilisation of sequencing data information in the HLA region, thereby reducing the false positive rate of HLA genotyping.

### HLA Genotyping

The HLA-HD [60] software (https://www.genome.med.kyoto-u.ac.jp/HLA-HD/) is a high-fidelity tool designed for high-resolution and precision HLA genotyping. In our study, HLA genes were genotyped using the aforementioned software, with all reads that were potentially derived from the HLA genes realigned to the reference sequences from the IMGT/HLA database (Release 3.45.0). To evaluate the accuracy of HLA genotyping, we employed the SBT-PCR-based “Golden Sets” HLA typing data from 1206 individuals from the 1KGP [7] (https://ftp.1000genomes.ebi.ac.uk/vol1/ftp/technical/working/20140725_hla_genotyp <u>es/</u>). The accuracy at each HLA gene is calculated by summing across the dosage of each correctly inferred HLA allele across all individuals (*n*), and divided by the total number of observations (2*n*) [13]. That is,

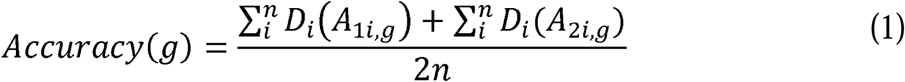

where *Accuracy(g)* represents the accuracy at a classical HLA gene (for example, *HLA-C*). *D_i_* represents the inferred dosage of an allele in individual *i*, and alleles *A_li,g_* and *A_2i,g_* represent the true (SBT-PCR-based “Golden Sets”) HLA types for an individual *i*.

### HLA-peptides Binding Affinity Analysis

In our investigation of the adaptability of HLA alleles to various epidemic pathogens (Table S1), we established a quantitative measure to define the affinity between an HLA molecule and a pathogen antigen as follows:

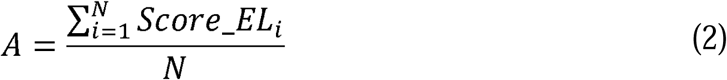

In this equation, *N* denotes the total count of peptides derived from the antigen.

The term *Score_EL_i_* signifies the predicted binding score (Predicted by NetMHCpan-4.1 and NetMHCIIpan-4.0 [61]) for the HLA molecule and the peptide that has significant binding affinity, comprising strong binders if the binding affinity ranking is in the top 0.5% and weak binders if the binding affinity ranking is in the top 2%. The quantification of HLA-peptide binding affinity is a well-established approach for prognosing the pathogen resistance of individual hosts [62]. Consequently, the metric *A* serves as a descriptive tool to articulate the likelihood of a host’s immune system effectively identifying and responding to pathogenic threats.

### Correlation Analysis of HLA-peptides Affinity and Pathogen Resistance

In order to evaluate the relationship between HLA-peptide binding affinity and pathogen resistance, we employed the mean value of predicted binding scores for HLA interactions with a pathogen’s peptides as an evaluative metric. In order to gain insight into the clinical adaptation of HIV and HCV infections in relation to the HLA genotypes of their hosts, we computed the predicted mean binding scores for the engagement of HLA molecules with pathogenic peptides, drawing on two studies that had previously explored this topic. Subsequently, we compared these scores with various measures of pathogen resistance, including viral setpoint, progression of viral disease, and the status of viral RNA level, in order to ascertain the efficacy of HLA genes across different levels of binding affinity.

### Reconstruction of HLA Reference Panel

Following the established pipeline for constructing an HLA reference panel (https://github.com/immunogenomics/HLA-TAPAS) [13], we integrated HLA genotypes and haplotype reference panel that were delineated from SNPs and indels within the HLA region. The initial step entailed the recapturing of variants that had been absent from the initial analysis due to the high degree of polymorphism characteristic of HLA genes. This was accomplished by realigning the sequences of the genotyped HLA genes to the GRCh38 assembly. Subsequently, variants with a minor allele frequency exceeding 0.001 within the MHC region were extracted from the haplotype reference panels of the NyuWa Han Chinese, the 1KGP, and the HGDP. In the final step, the HLA genotypes were integrated into the aforementioned haplotype reference panels, after which the haplotypes were re-phased using Shapeit4 [63].

### Selection Analysis of HLA Regulators

To detect adaptive selection within HLA regulatory regions, we systematically computed the single density score (SDS), integrated haplotype score (iHS) statistics for genome-wide detection of recent positive selections and Beta statistics for long-term balancing selections in the NyuWa Han Chinese population [64–67]. In order to ensure the reliability of the results, we applied rigorous criteria to common variants, defined by a minor allele frequency (MAF) of 0.05 or greater. Subsequently, the aforementioned common variants were employed in the calculation of SDS, iHS and Beta statistics. Statistics with a false discover rate (FDR) adjusted using the Benjamini-Hochberg procedure of less than 0.05 (|SDS| >= 3.857; |iHS| >= 4.234; Beta >= 8.41) are considered to indicate significant selection signals. To ascertain the robustness of potential selective sweeps or balancing selection events, a sliding window approach was employed, with each segment comprising 100 SNPs and a step size of 50 SNPs. Intervals with a higher proportion of significant signals within this framework were considered as indicative of more robust selective events. To substantiate the implications of these selection signals on phenotypic variation, we leveraged eQTL data from the GTEx project, which provided evidence for alterations in gene expression phenotypes at the transcriptional level [68]. The integration of selection statistics with functional genomics data enabled the potential fitness consequences of adaptive regulatory changes in the HLA region to be inferred.

### Allele Frequency Trajectories Computation

An approximate full-likelihood method [69] was employed to deduce the selection coefficients and allele frequency trajectories for the alleles under selection. In this analysis, our focus was on a 5kb haploblock, characterised by infrequent recombination, which encompasses the two adaptive sites of interest. This region was selected for the reconstruction of allele frequency trajectories over the last 1000 generations, with the stipulation that the insertions and deletions within the haploblock were not considered. In preparation for inferring the allele frequency trajectories, the Relate software [70] was employed to estimate the genealogical and population history of the haploblock. For the Han Chinese population, the effective population size was set to 20,000, the mutation rate to 1.25e-8, the years per generation to 28, and the number of sampling times of branch lengths to 100. All other parameters were configured according to their default settings.

## Ethical statement

This study was approved by the Medical Research Ethics Committee of Institute of Biophysics, Chinese Academy of Sciences. All participants provided written informed consent. The informed consent is used to collect samples for genome studies conducted by Chinese Academy of Sciences. The consent requires participants to be 30-70 years old patients and healthy people with full capacity. Participants voluntarily donate blood samples, provide clinical treatment information and sign informed consent. All their personal information is kept confidential. Participants can choose not to participate in sample donation, or withdraw at any time.

## Data Availability

The DNA sequencing data of NyuWa samples used in this study have been deposited in the Genome Sequence Archive (GSA) in National Genomics Data Centre, China National Centre for Bioinformation/Beijing Institute of Genomics, Chinese Academy of Sciences, under accession number HRA004185 (https://ngdc.cncb.ac.cn/gsa-human/). These data are available under restricted access for privacy protection and can be obtained by application on the GSA database website following the guidance of “Request Data” on this website. These data have also been deposited in the National Omics Data Encyclopaedia (NODE) of the Bio-Med Big Data Centre, Shanghai Institute of Nutrition and Health, Chinese Academy of Sciences, under accession number OEP002803 (http://www.biosino.org/node). The user can register and login to this website and follow the guidance of “Request for Restricted Data” to request the data. The reference genome GRCh38 used in this study is available at https://console.cloud.google.com/storage/browser/genomicspublic-data/resources/broad/hg38/v0/.

## CRediT authorship contribution statement

**Shuai Liu:** Conceptualization, Investigation, Methodology, Formal analysis, Visualization, Writing – original draft, Writing – review & editing. **Yanyan Li:** Conceptualization, Investigation, Methodology, Formal analysis, Visualization, Writing – original draft, Writing – review & editing. **Tingrui Song:** Conceptualization, Methodology, Formal analysis, Visualization, Writing – original draft, Writing – review & editing. **Jingjing Zhang:** Investigation, Methodology, Formal analysis. **Peng Zhang:** Data curation, Methodology, Formal analysis. **Huaxia Luo:** Methodology, Formal analysis. **Sijia Zhang:** Data curation. **Yiwei Niu:** Data curation. **Tao Xu:** Conceptualization, Funding acquisition, Project administration. **Shunmin He:** Conceptualization, Funding acquisition, Project administration, Writing – review & editing. All authors have read and approved the final manuscript.

## Acknowledgements

We thank Zhen Xiong for thoughtful discussions and valuable comments regarding the immunogenetics. We thank the people for generously contributing samples to the NyuWa dataset. Data analysis and computing resources were supported by the Centre for Big Data Research in Health (http://bigdata.ibp.ac.cn), Institute of Biophysics, Chinese Academy of Sciences. This work was supported by Strategic Priority Research Program of the Chinese Academy of Sciences [XDB38040300]; National Key R&D Program of China [2021YFF0703701, 2021YFF0704500, 2022YFC3400405]; 14th Five-year Informatization Plan of Chinese Academy of Sciences [CAS-WX2021SF-0203]; National Natural Science Foundation of China [91940306, 31970647, 32200478]; China Postdoctoral Science Foundation [2022M713311, GZC20232899]; and National Genomics Data Centre, China.

## Declaration of AI and AI-assisted technologies in the writing process

In the writing process section, we did not use the AI and AI-assisted technologies for generating content or images, writing code and processing data, and only apply them to the use of basic tools for checking grammar, spelling.

