## Supplementary material for "The Pathogen Adaptation of HLA Alleles and the Correlation with Autoimmune Diseases based on the HLA Diversity Resource in the Han Chinese": Figure S: Supplementary_materials.docx

Conflict of interest declaration: We declare we have no competing interests.

**Table S1. Pathogenic antigen data information.**

| **Categories** | **Pathogens** | **Antigens (NCBI ID or UniProt ID)** |
| --- | --- | --- |
| Intracellular | *Hepatitis C virus* | UP000000518 |
| Intracellular | *Rubella virus* | UP000000571 |
| Intracellular | *Variola virus* | UP000002060 |
| Intracellular | *HIV1* | UP000002241 |
| Intracellular | *Mumps virus* | UP000002331 |
| Intracellular | *Hepatitis B virus* | UP000007930 |
| Intracellular | *Rabies virus* | UP000008649 |
| Intracellular | *Measles virus* | UP000008699 |
| Intracellular | *Influenza A virus* | UP000009255 |
| Intracellular | *Epstein-Barr virus* | UP000153037 |
| Intracellular | *MERS-CoV* | UP000171868 |
| Intracellular | *SARS-CoV* | UP000000354 |
| Intracellular | *SARS-CoV2* | UP000464024 |
| Intracellular-Extracellular | *Treponema pallidum* | AAA75016, AAA27481, AAA27477, AAA27472, AAC45732 |
| Intracellular-Extracellular | *Mycobacterium leprae* | NP_302372, NP_301968, NP_301879, NP_301372, CAR70980 |
| Intracellular-Extracellular | *Toxoplasma gondii* | EPT30382, EPT30138, EPT30276, EPT29989, EPT26499, EPT27242, EPT26403, EPT30357, EPT29845 |
| Intracellular-Extracellular | *Mycobacterium tuberculosis* | NP_218391, YP_177853, YP_177893, NP_216548, NP_218393, YP_178022, NP_215554, NP_218321, NP_214989, NP_216495 |
| Intracellular-Extracellular | *Streptococcus pneumoniae* | NP_357715, NP_359586, NP_359346, NP_358461, NP_359024, NP_358176, NP_359129, NP_359087, NP_358155, NP_358175 |
| Intracellular-Extracellular | *Yersinia pestis* | NP_395430, NP_395429, NP_395427, NP_395165, NP_395166, NP_395143 |
| Intracellular-Extracellular | *Bordetella pertussis* | NP_882282, NP_882283, NP_882286, NP_882284, NP_882285, NP_879898, NP_880302, NP_880571, NP_879839, NP_881965 |
| Intracellular-Extracellular | *Corynebacterium diphtheriae* | NP_938615 |
| Intracellular-Extracellular | *Salmonella enterica* | NP_456106, NP_456107, NP_456109, NP_456110, NP_456114, NP_456134, NP_456135, NP_456136, NP_458730, NP_458731, NP_458734, NP_458732, NP_458733, NP_458738, NP_458739, NP_458741, NP_458735 |
| Intracellular-Extracellular | *Plasmodium falciparum* | XP_001348275, XP_001348247, XP_001350083, XP_002809051, XP_001349749, XP_001350088, XP_001349859, XP_001350569, XP_001350410, XP_001348153, XP_001348015, XP_001347895, XP_001347636, XP_001347630, XP_001347629, XP_001352222, XP_001352170, XP_001349336, XP_002808637, XP_001349578 |
| Extracellular | *Clostridium tetani* | NP_783831, YP_008774065, NP_782184, NP_780878, NP_781182 |
| Extracellular | *Vibrio cholera* | NP_231099, NP_231100, NP_231102, NP_231104 |
| Extracellular | *Bacillus anthracis* | UP000000594 |
| Extracellular | *Entamoeba histolytica* | XP_655241, EAL50306, EAL50995, XP_652992, AAC72364, XP_657050, XP_649161, AAA29100, XP_650725, XP_648032 |
| Extracellular | *Giardia lamblia* | XP_001704065, XP_001705852, XP_001705828, XP_001703888, XP_001705844, XP_001705829, XP_001705785, XP_001705784, XP_001705783, XP_001703933, XP_001703925, XP_001703844, XP_001703931 |
| Extracellular | *Trichinella spiralis* | CAA73574, EFV52545, ACV51809, EFV53657 |
| Extracellular | *Trichomonas vaginalis* | XP_001292151, XP_001313891, XP_001284871, XP_001580136, XP_001582296, XP_001299798, XP_001584317, XP_001327241, XP_001325298, XP_001303631, XP_001316801, XP_001326883, XP_001301868 |
| Extracellular | *Schistosoma mansoni* | CCD74732, CCD75328, CCD75626, CCD75625, CCD75627, CCD80234, CCD77656, CCD77655 |

**Table S2. Autoimmune diseases related data information.**

| Traits | Related Genes | Types | References |
| --- | --- | --- | --- |
| Multiple sclerosis | HLA-DRB1*15 | risk | [1] |
| Multiple sclerosis | HLA-DQB1*06 | risk | [1] |
| Multiple sclerosis | HLA-B*03:01 | risk | [2] |
| Multiple sclerosis | HLA-DRB1*13:03 | risk | [2] |
| Multiple sclerosis | HLA-DRB1*03:01 | risk | [2] |
| Multiple sclerosis | HLA-DRB1*08:01 | risk | [2] |
| Multiple sclerosis | HLA-DQA1*01:02 | risk | [2] |
| Multiple sclerosis | HLA-DQB1*06:02 | risk | [2] |
| Multiple sclerosis | HLA-DQB1*03:02 | risk | [2] |
| Multiple sclerosis | HLA-A*02:01 | protective | [2] |
| Rheumatoid arthritis | HLA-DRB1*04:01 | risk | [3] |
| Rheumatoid arthritis | HLA-DRB1*04:04 | risk | [3] |
| Rheumatoid arthritis | HLA-DRB1*04:05 | risk | [3] |
| Rheumatoid arthritis | HLA-DRB1*04:10 | risk | [3] |
| Rheumatoid arthritis | HLA-DRB1*04 | risk | [4] |
| Rheumatoid arthritis | HLA-DRB1*13:01 | protective | [5] |
| Rheumatoid arthritis | HLA-DRB1*13:02 | protective | [5] |
| Rheumatoid arthritis | HLA-DQB1*06:01 | risk | [5] |
| Coeliac disease | HLA-DQB1*02:01 | risk | [6] |
| Coeliac disease | HLA-DQB1*02:02 | risk | [6] |
| Coeliac disease | HLA-DQB1*03:02 | risk | [6,7] |
| Coeliac disease | HLA-DQB1*03:05 | risk | [7] |
| Coeliac disease | HLA-DQA1*05:01 | risk | [8] |
| Inflammatory bowel diseases | HLA-A*03:01 | risk | [9] |
| Inflammatory bowel diseases | HLA-C*06:02 | risk | [9] |
| Inflammatory bowel diseases | HLA-C*08:02 | risk | [9] |
| Inflammatory bowel diseases | HLA-C*12:02 | risk | [9] |
| Inflammatory bowel diseases | HLA-C*14:02 | risk | [9] |
| Inflammatory bowel diseases | HLA-B*08:01 | protective | [9] |
| Inflammatory bowel diseases | HLA-B*57:01 | risk | [9] |
| Inflammatory bowel diseases | HLA-B*14:02 | risk | [9] |
| Inflammatory bowel diseases | HLA-B*35:03 | protective | [9] |
| Inflammatory bowel diseases | HLA-B*52:01 | risk | [9] |
| Inflammatory bowel diseases | HLA-B*35:02 | risk | [9] |
| Inflammatory bowel diseases | HLA-DRB1*01:03 | risk | [9] |
| Inflammatory bowel diseases | HLA-DRB1*01:01 | protective | [9] |
| Inflammatory bowel diseases | HLA-DRB1*03:01 | protective | [9] |
| Inflammatory bowel diseases | HLA-DRB1*07:01 | risk | [9] |
| Inflammatory bowel diseases | HLA-DRB1*08:01 | risk | [9] |
| Inflammatory bowel diseases | HLA-DRB1*16:01 | protective | [9] |
| Inflammatory bowel diseases | HLA-DRB1*13:02 | risk | [9] |
| Inflammatory bowel diseases | HLA-DQA1*02:01 | risk | [9] |
| Inflammatory bowel diseases | HLA-DQA1*04:01 | risk | [9] |
| Inflammatory bowel diseases | HLA-DQB1*02:01 | protective | [9] |
| Inflammatory bowel diseases | HLA-DQB1*04:02 | risk | [9] |
| Inflammatory bowel diseases | HLA-DQB1*05:02 | protective | [9] |
| Inflammatory bowel diseases | HLA-DPA1*01:03 | protective | [9] |
| Psoriasis | HLA-C*06 | risk | [10] |
| Type 1 diabetes | HLA-DRB1*03 | risk | [11] |
| Type 1 diabetes | HLA-DRB1*04 | risk | [11] |
| Type 1 diabetes | HLA-DQB1*03:02 | risk | [12] |
| Type 1 diabetes | HLA-DQB1*02:01 | risk | [12] |
| Type 1 diabetes | HLA-DQA1*03:01 | risk | [12] |
| Type 1 diabetes | HLA-DQA1*01:01 | protective | [12] |
| Type 1 diabetes | HLA-DQB1*05:03 | protective | [12] |
| Type 1 diabetes | HLA-DQB1*06:02 | protective | [12] |
| Type 1 diabetes | HLA-DQB1*06:03 | protective | [12] |
| Type 1 diabetes | HLA-DQB1*06:04 | protective | [12] |
| Type 1 diabetes | HLA-DRB1*09:01 | risk | [13] |
| Type 1 diabetes | HLA-DRB1*04:05 | risk | [13] |
| Type 1 diabetes | HLA-DRB1*15:03 | protective | [13] |

**Figure S1. Major HLA Alleles in the Han Chinese. A.** NyuWa Han Chinese. **B.** East Asian. **C.** European. **D.** African.

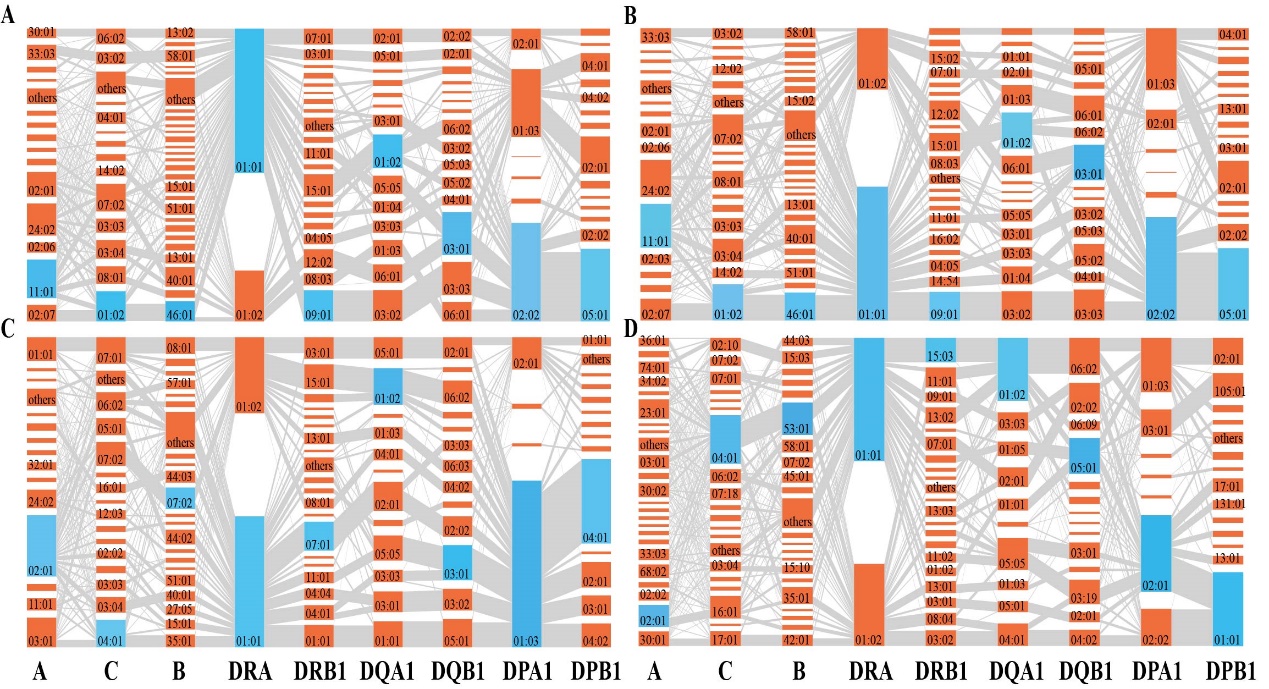

**Figure S2. Relationship between HIV-associated HLA genes and HIV invasion ability and in vivo reproductive ability. Viral setpoint:** the time at which plasma viraemia settles to a relatively stable level (within approximately 3–6 months of the onset of HIV infection). Viral setpoint is strongly predictive of both how quickly HIV infection will progress and the risk of HIV transmission[14]**. Progression:** the rate of disease progression[14]. **Scaled score EL**: the mean of binding affinity of HLA molecule with peptides of a pathogen.

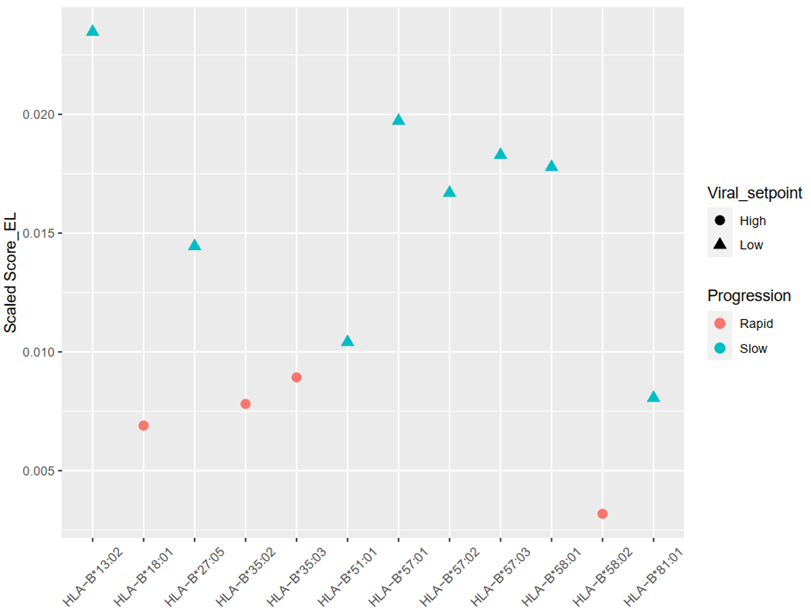

**Figure S3. Relationship between HCV-associated HLA genes and the RNA status of HCV. HCV RNA status:** HCV RNA level in vivo.

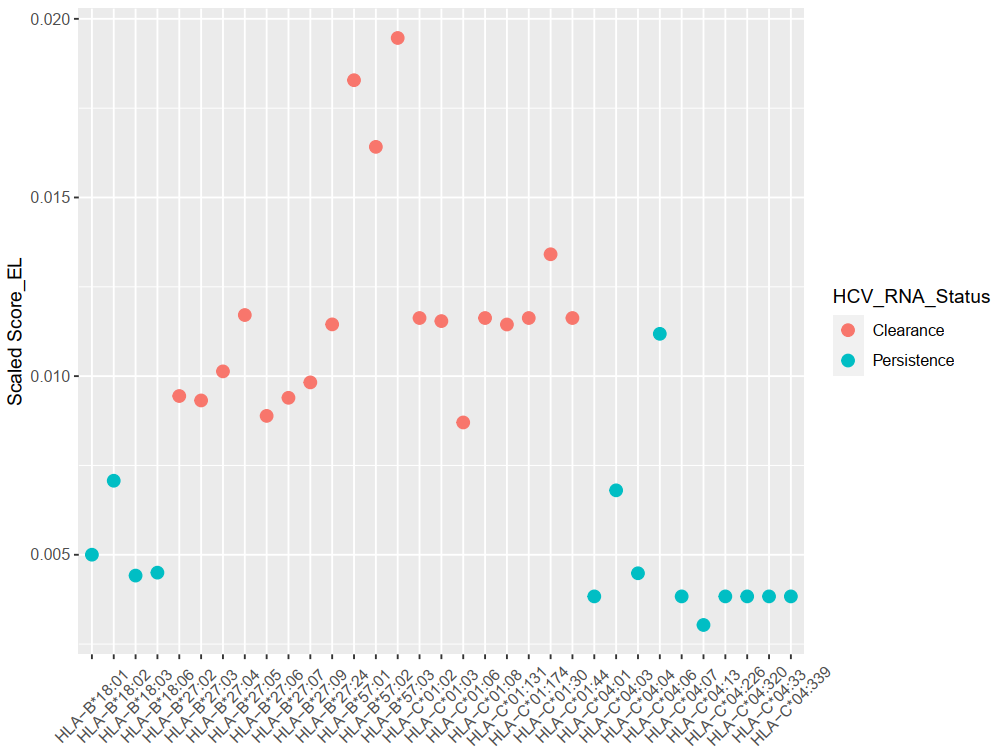

**Figure S4. The binding affinity between the epidemic pathogens and the rare HLA types in the Han Chinese population.** The pathogens in the yellow block are virus, the pathogens in the pink block are bacteria, and the pathogens in the blue block are parasites. The higher score means higher affinity.

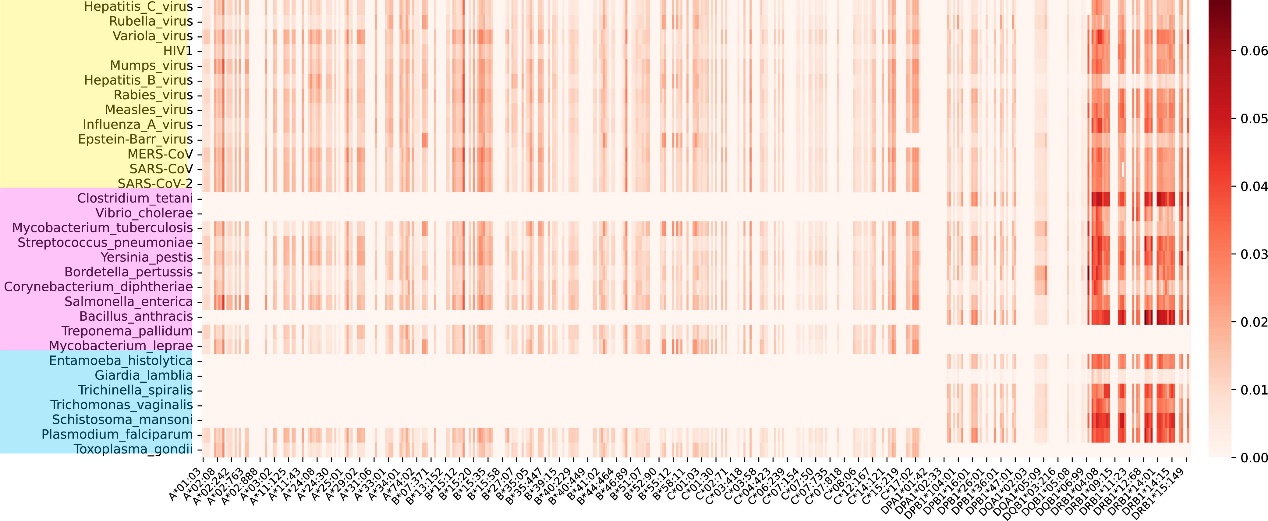

**Figure S5. The binding affinity between the epidemic pathogens and the common HLA types in the European population and the African population. A.** European**. B.** African. The pathogens in the yellow block are virus, the pathogens in the pink block are bacteria, and the pathogens in the blue block are parasites. The higher score means higher affinity.

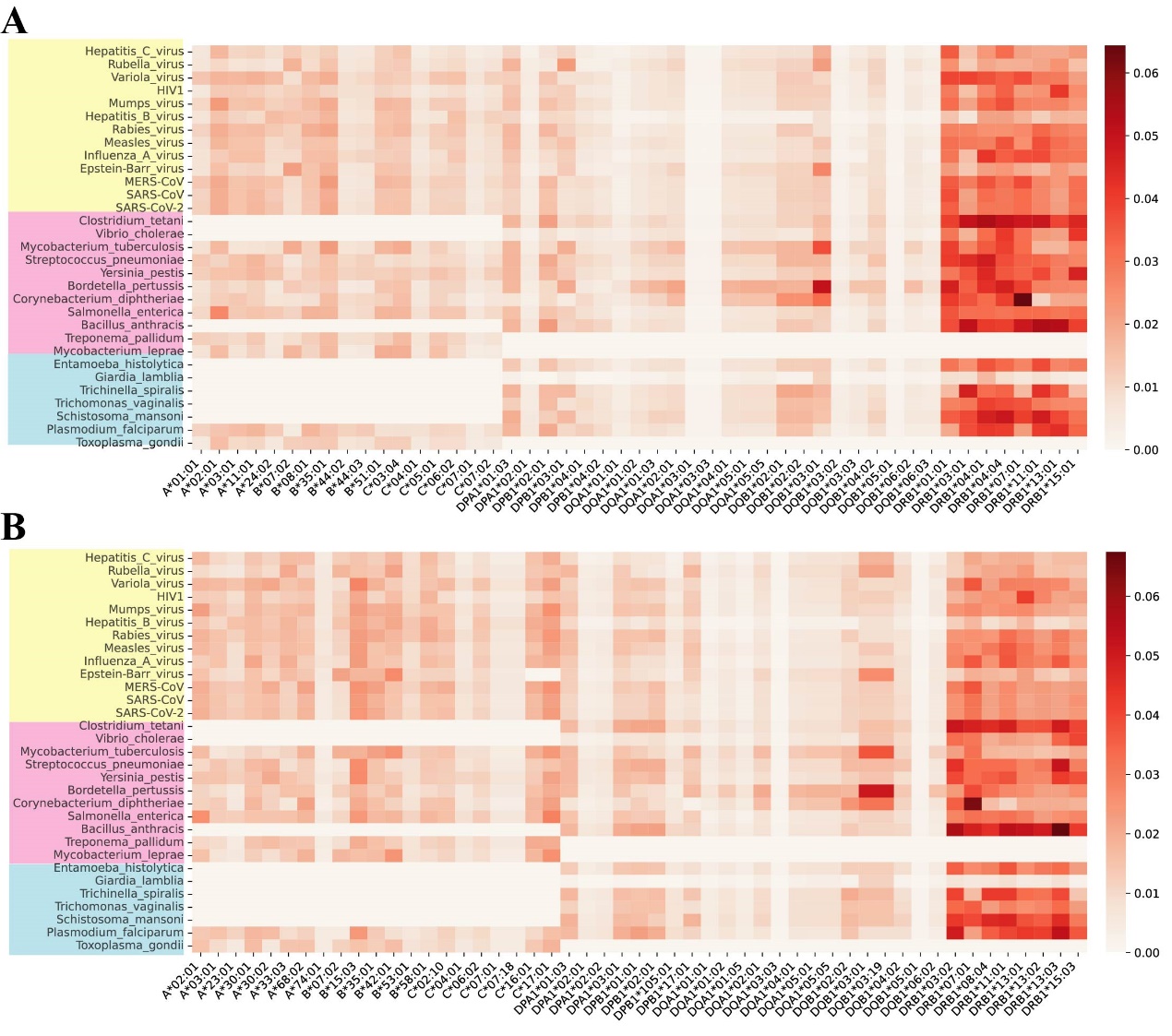

**Figure S6. The binding affinity between the epidemic pathogens and the low frequency and rare HLA types in the European population and the African population. A.** European**. B.** African. The pathogens in the yellow block are virus, the pathogens in the pink block are bacteria, and the pathogens in the blue block are parasites. The higher score means higher affinity.

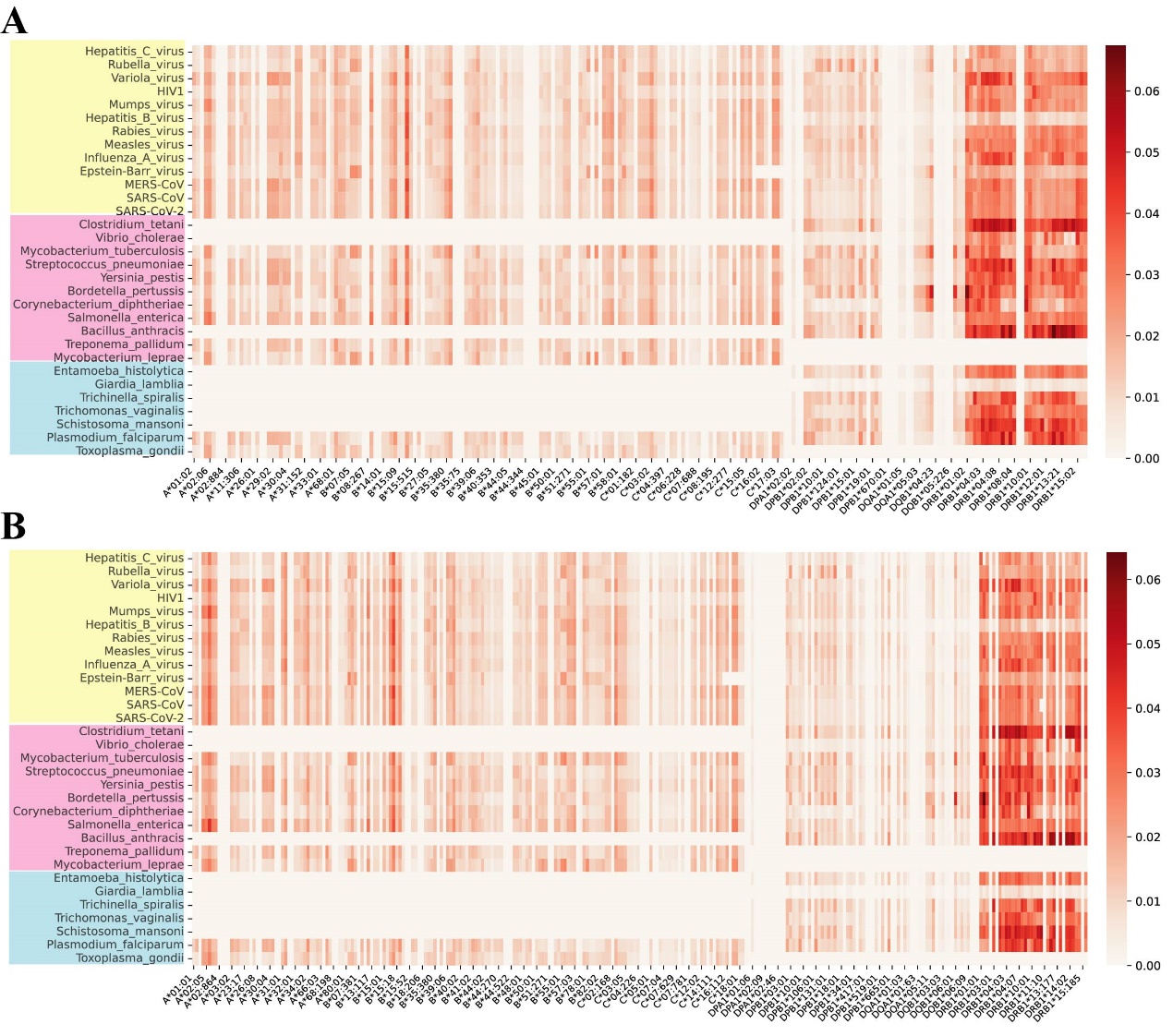

**Figure S7. Affinity preference of three *HLA-DRB1* alleles for pathogen peptides. A.** Peptide-binding preference of HLA-DRB1*07:01 for *Corynebacterium diphtheriae*. **B.** Peptide-binding preference of HLA-DRB1*08:03 for *Clostridium tetani*. **C.** Peptide-binding preference of HLA-DRB1*08:03 for *Bacillus anthracis*. **D.** Peptide-binding preference of HLA-DRB1*14:54 for *Bacillus anthracis*.

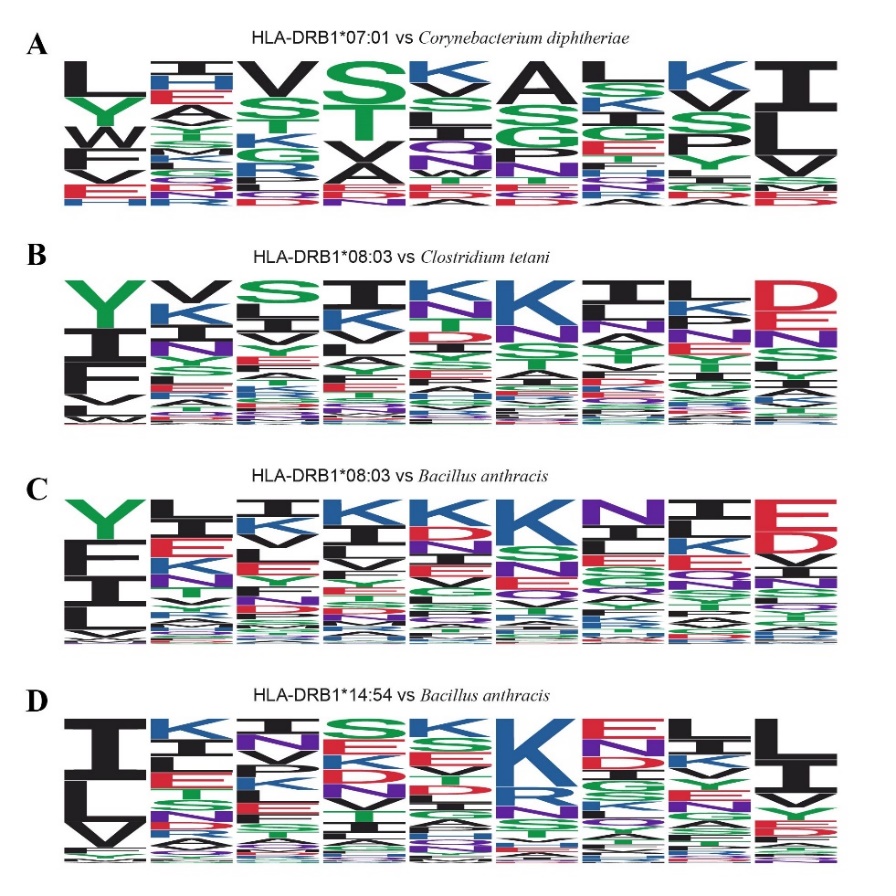

**Figure S8. Affinity preference of two *HLA-DQB1* alleles for pathogen peptides. A.** HLA-DQB1*03:01. **B.** HLA-DQB1*06:01. Here mainly revolves 3 pathogens, including *Bordetella pertussis*, *Mycobacterium tuberculosis*, *Corynebacterium diphtheriae*.

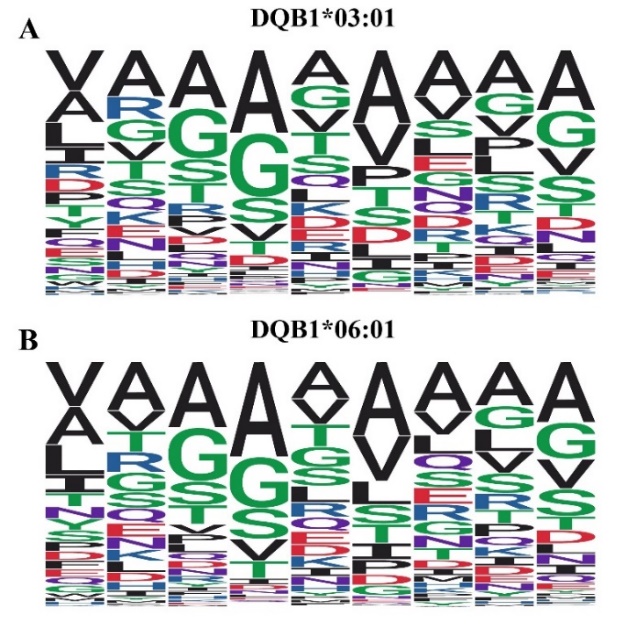

**Figure S9. Correlation between the low-frequency HLA alleles in the Han Chinese and the autoimmune susceptibility HLA alleles.** The horizontal axis represents common potential adaptive HLA alleles in the Han Chinese population, and the vertical axis represents autoimmune susceptibility HLA alleles. Pearson correlation tests were performed on the genotypes of the two sets of gene sets, “*” indicates significantly correlated gene pairs (r^2^>0.2 and P<0.05).

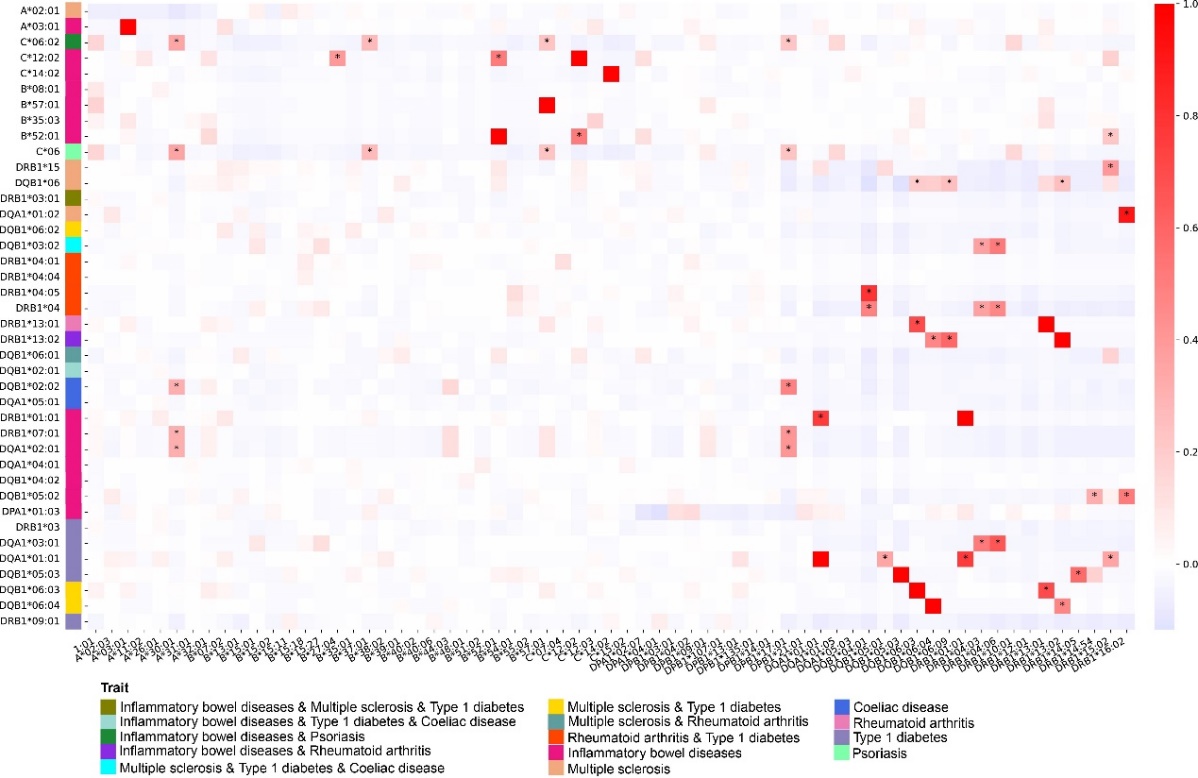

**Figure S10. Correlation between the common HLA alleles and the autoimmune susceptibility HLA alleles in the European population and the African population. A.** European**. B.** African. The horizontal axis represents common potential adaptive HLA alleles in the Han Chinese population, and the vertical axis represents autoimmune susceptibility HLA alleles. Pearson correlation tests were performed on the genotypes of the two sets of gene sets, “*” indicates significantly correlated gene pairs (r^2^>0.2 and P<0.05).

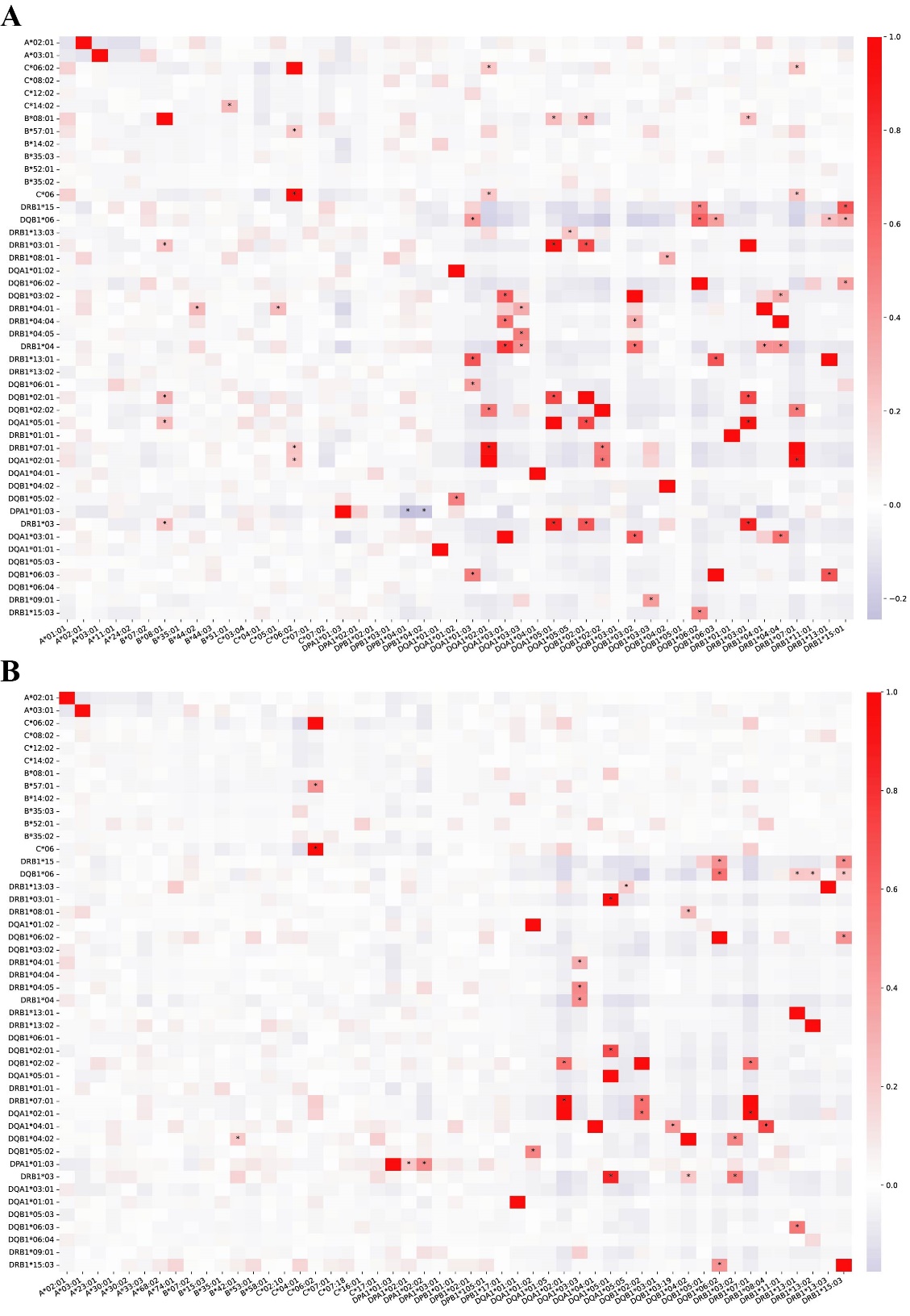

**Figure S11. Correlation between the low frequency and rare HLA alleles and the autoimmune susceptibility HLA alleles in the European population and the African population. A.** European**. B.** African. The horizontal axis represents common potential adaptive HLA alleles in the Han Chinese population, and the vertical axis represents autoimmune susceptibility HLA alleles. Pearson correlation tests were performed on the genotypes of the two sets of gene sets, “*” indicates significantly correlated gene pairs (r^2^>0.2 and P<0.05).

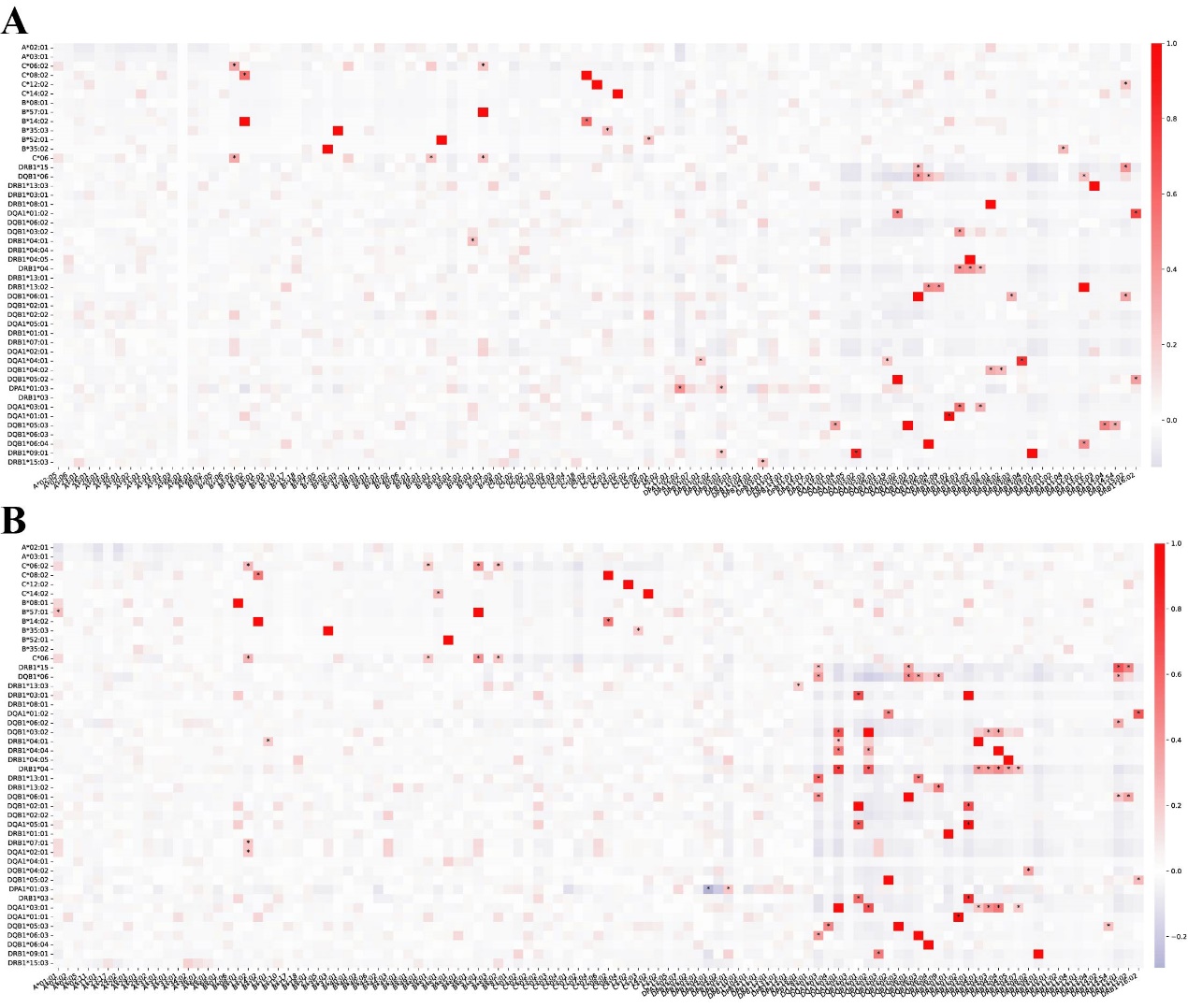

**Figure S12. The adaptive selection signals in the MHC region.** The dashed line marked the significant level of statistics.

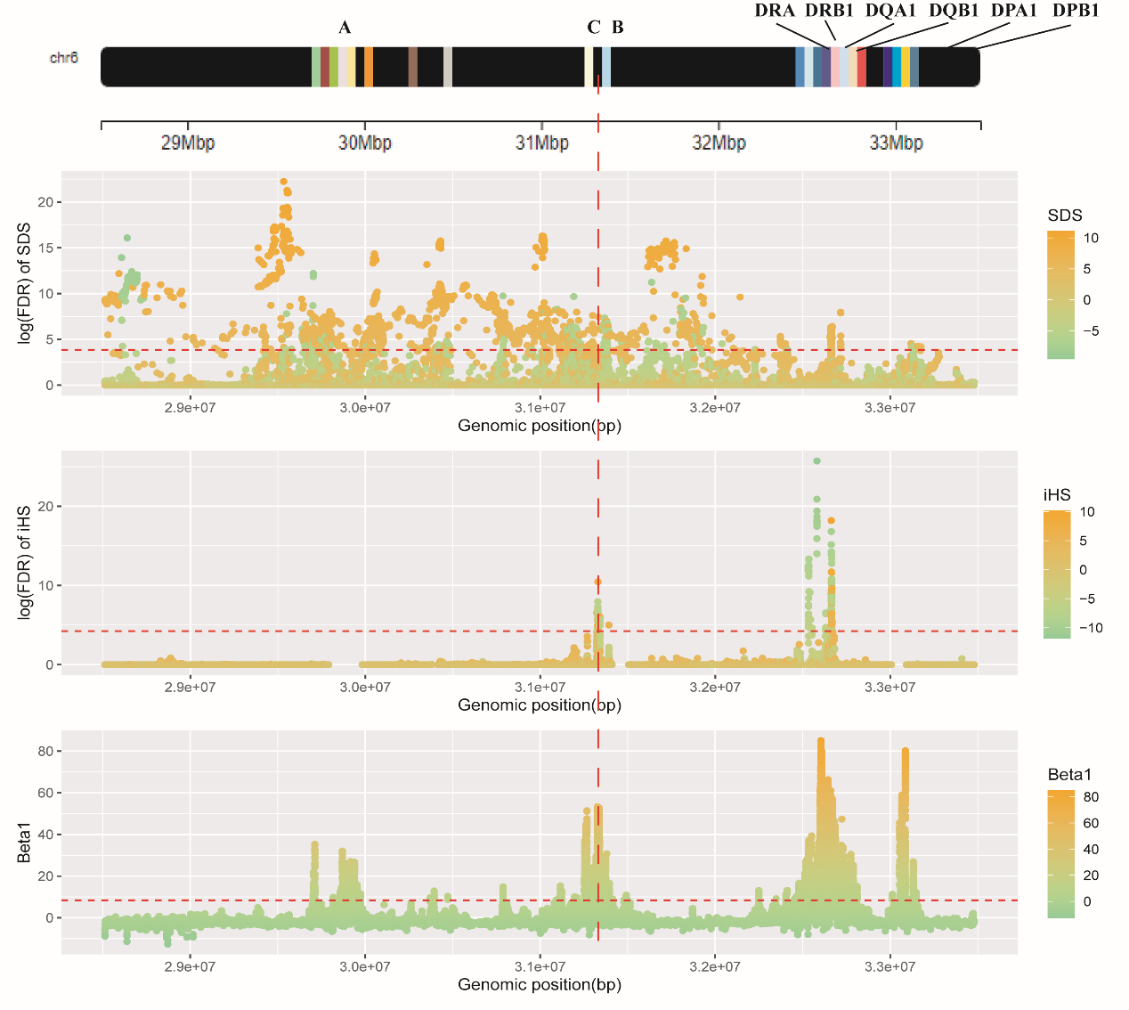

**Figure S13. The adaptive selection signals in 6p21.33. A.** genetic diversity. **B.** Recombination rate. **C.** Proportion of beta1 greater than 8.41 in a 100 snp window. **D.** Proportion of |iHS| greater than 4.234 in a 100 snp window.

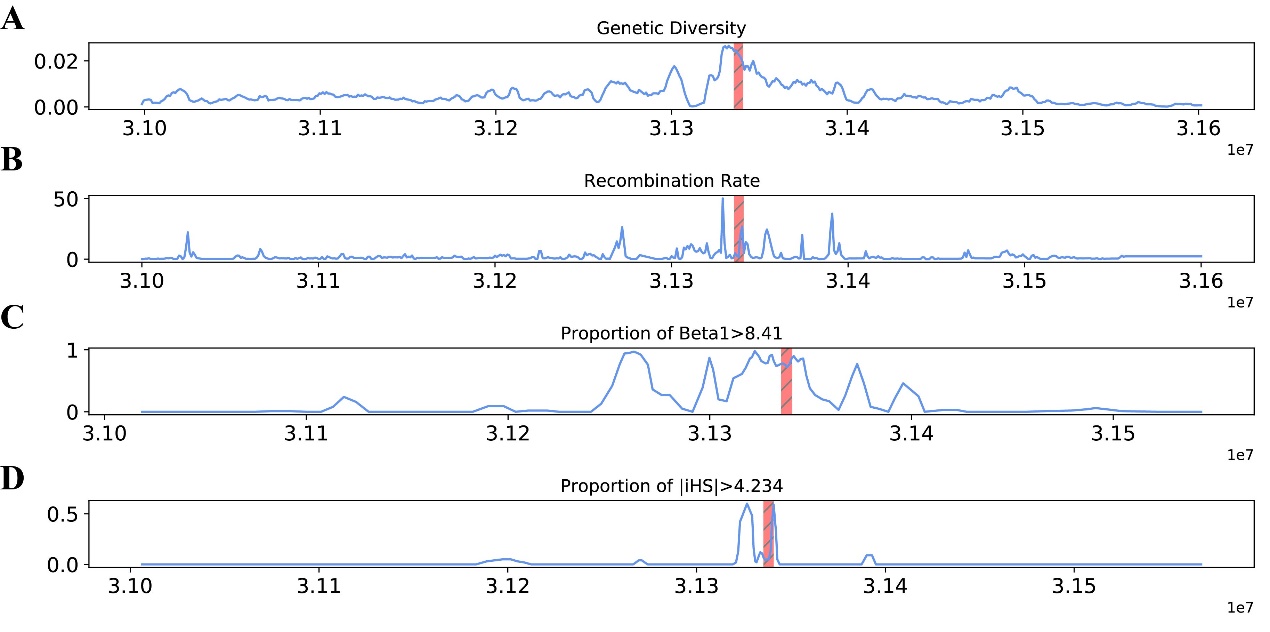

**Figure S14. The allele frequencies of the adaptive variants in East Asian, European and African from 1KGP.**

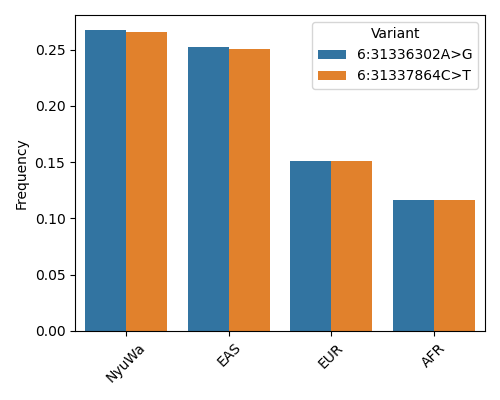

**Figure S15. Allele frequency trajectories of 6:31336302G in recent 1000 generations.**

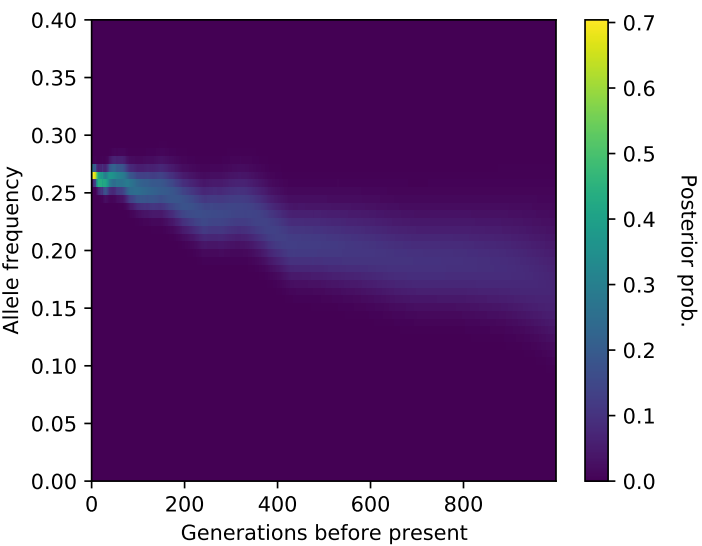

**Figure S16. Allele frequency trajectories of 6:31337864T in recent 1000 generations.**

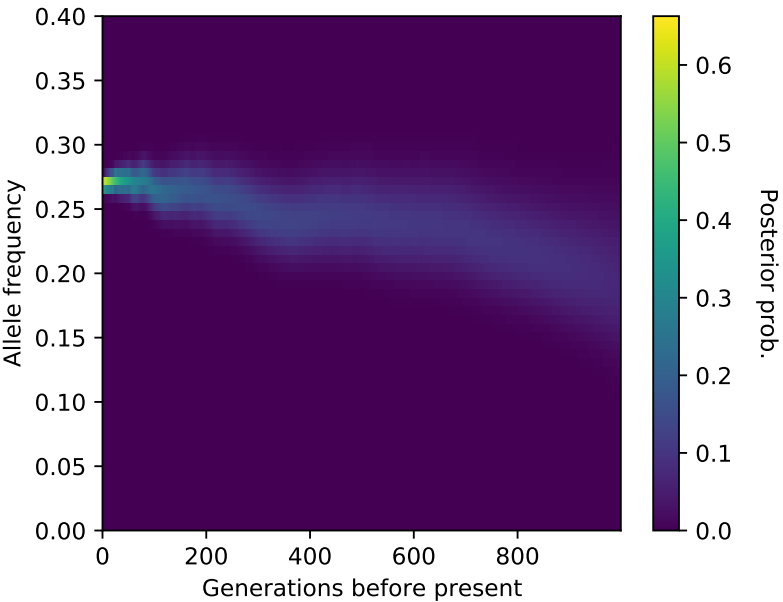

**Figure S17. Affinity preference of HLA-C*03:02 for all pathogen peptides. A.** Sequence characteristics of all pathogen antigen peptides that have the ability to bind to HLA-C*03:02. **B.** Sequence characteristics of all pathogen antigen peptides with strong binding ability to HLA-C*03:02. **C.** Sequence characteristics of all pathogen antigen peptides with weak binding ability to HLA-C*03:02.

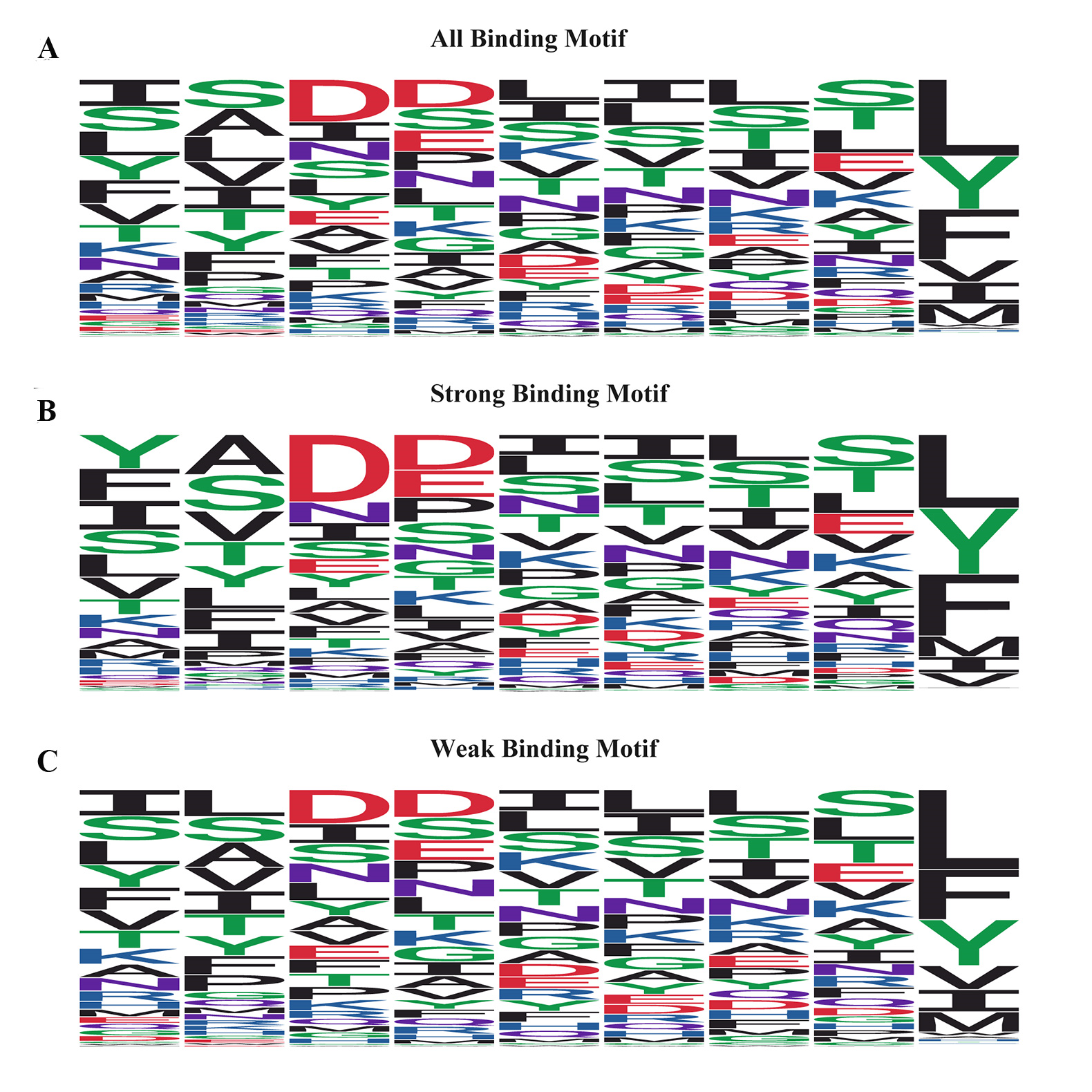
